# Gene-diet interaction analysis in UK Biobank identified genetic loci that modify the association between fish oil supplementation and the incidence of dementia

**DOI:** 10.1101/2024.11.08.24316999

**Authors:** Yueqi Lu, Huifang Xu, Yitang Sun, Susan Adanna Ihejirika, Charleston W. K. Chiang, Burcu F. Darst, Suhang Song, Ye Shen, Kaixiong Ye

**Affiliations:** Department of Genetics, University of Georgia, Athens, Georgia, US; Institute of Bioinformatics, University of Georgia, Athens, Georgia, US; Center for Genetic Epidemiology, Department of Population and Public Health Sciences, University of Southern California, Los Angeles, California, US; Public Health Sciences, Fred Hutchinson Cancer Center, Seattle, Washington, US; Department of Epidemiology, University of Washington, Seattle, Washington, US; Department of Health Policy and Management, College of Public Health, University of Georgia, Athens, Georgia, US; Department of Epidemiology and Biostatistics, College of Public Health, University of Georgia, Athens, Georgia, US

**Keywords:** Time-to-event GWAS, gene-by-environment analysis, fish oil supplementation, dementia

## Abstract

**Background:** Dementia is a common disease influenced by both genetic and environmental factors. *APOE* ε4 is well-known to increase the risk of dementia, and it has been shown to attenuate the protective association of fish oil supplementation and the incidence of dementia. To identify more genetic factors with similar modifying effects, we performed a genome-wide scan.

**Methods:** We first performed time-to-event genome-wide association study (GWAS) of all-cause dementia and two of its subtypes, Alzheimer’s disease (AD) and vascular dementia, in the UK Biobank. GWAS were performed in all participants (N = 357,631) and in two subgroups with or without fish oil supplementation (N = 113,267 and 244,364, respectively). Single nucleotide polymorphisms (SNPs) suggestively associated with dementia were then evaluated for their interactions with fish oil status in Cox-regression models. Furthermore, we conducted gene set enrichment analysis to identify the relevant cell types for these interaction signals.

**Results:** Time-to-event GWAS identified 6, 5, and 2 genome-wide significant loci (p < 5e-8) for the incidence of all-cause dementia, AD, and vascular dementia, respectively. Most of them overlapped with previously known GWAS loci for AD and related dementia. A total of 178 suggestive GWAS loci (p < 1e-5) were passed onto interaction analysis, and 43 of them were found to significantly modify the association between fish oil supplementation and dementia incidence (p < 2.8e-4 with Bonferroni correction). One locus overlapped with a known AD GWAS locus (*EED*/*PICALM*) and two overlapped with GWAS loci for circulating omega-3 fatty acids (*SRSF4*, *PSMG1*). Gene set enrichment analysis found that candidate genes of interaction signals demonstrated tissue or cell-type specificity in the brain.

**Conclusion:** We identified 43 genetic loci that modify the association between fish oil supplementation and dementia. These findings indicate a need for genome-informed personalized nutrition of fish oil supplementation for the purpose of dementia prevention.

## Introduction

Dementia is a significant global health issue exacerbated by population aging and growth.^1^ This syndrome, characterized by the loss of cognitive functioning, is influenced by both genetic and environmental factors.^2–4^ Nutrition and dietary components are considered as the modifiable risk factors of dementia.^2^ Observational studies reported that omega-3 fatty acids were associated with a lower risk of cognitive decline.^5^ Habitual intake of fish oil supplements, a rich source of long-chain omega-3 fatty acids, was found to be inversely associated with the risk of all-cause dementia and several subtypes.^6^ The impacts of genetic factors have been examined by multiple genome-wide association studies (GWAS).^4,7,8^ A meta-study in 2023 revealed more than 75 loci associated with incident Alzheimer’s disease and related dementias (ADRD).^4^ One notable gene is *APOE*, with its risk allele *APOE* ε4 linked to vascular dysfunction, amyloid-β pathology, neurodegeneration, and ultimately, dementia.^9–13^

Gene-environment (G×E) interaction studies on human health are gaining widespread attention. These studies focus on how specific genetic and environmental factors synergistically increase disease risk, providing new insights into the etiology of complex diseases and promoting the development of preventative strategies and therapies.^14^ Recent studies showed that *APOE* ε4 modifies the protective associations of fish oil supplementation with all-cause dementia and vascular dementia, suggesting that only non-carriers of *APOE* ε4 may benefit from fish oil supplementation.^15,16^ Besides *APOE*, the potential modifying effects of other genetic factors remain unknown.

In this study, we sought to identify additional genetic variants that modify the association of fish oil supplementation with all-cause dementia and its two main subtypes, Alzheimer’s disease and vascular dementia. Time-to-event models used throughout the analysis allowed the detection of genetic variants associated with the onset of diseases.^17^ We adopted a two-step approach by first identifying suggestive loci associated with dementia phenotypes in the overall sample and the two fish oil supplementation subgroups. Then, time-to-event interaction analysis was used to identify significant interaction loci with Bonferroni correction. Finally, we assessed the expression signatures of candidate genes at these interaction loci and identified their biologically relevant tissue or cell types.

## Methods

### Study design and participants

We included participants of genetic European ancestry from UK Biobank, who completed the touch-screen questionnaire for the fish oil supplementation, did not have a diagnosis of dementia at baseline, and did not withdraw from the study by April 25^th^, 2023. The ancestry background was obtained from the Pan-UK Biobank Project by projecting the UKB individuals onto the principal components (PCs) inferred from the reference panels of the Human Genome Diversity Panel (HGDP) and 1000 Genomes Project.^18^

### Outcomes

The dementia outcomes provided by UK Biobank were algorithmically defined using information from self-reported verbal interviews, linked hospital admissions, and death registries. Our study considered incident all-cause dementia, Alzheimer’s disease, and vascular dementia as the longitudinal outcomes. Incidence was defined as the first diagnosed date following the participant’s visit to the assessment center at baseline. The end of follow-up for each participant was recorded as the earliest of the following dates: dementia occurrence, death, or the end of follow-up records (Dec 13^th^, 2022).

### Exposure

The touch-screen questionnaire completed at the assessment center provided information on fish oil supplementation. Participants were asked, “Do you regularly take any of the following?”. Participants who selected the option “Fish oil (including cod liver oil)” were considered fish oil users, and those who only selected other supplementation or “None of the above” were considered non-fish oil users. To evaluate the robustness of this questionnaire, we performed a sensitivity analysis on fish oil intake status based on the 24-hour dietary recall questionnaire. More details of sensitivity analysis were described in Supplementary Methods.

### APOE ε4 dosage

*APOE* ε4 dosage was known to modify the relationship between fish oil supplementation and dementia onset.^15^ The *APOE* genotype was defined by two single nucleotide polymorphisms (SNPs), rs7412 and rs429358. The dosage of *APOE* ε4 was divided into 0 (ε1/ε1, ε1/ε2, ε2/ε2, ε2/ε3, ε3/ε3), 1 (ε1/ε4, ε3/ε4) and 2 (ε4/ε4), respectively. Since the determination of ε1/ε3 and ε2/ε4 from the genotype data was ambiguous, and ε2/ε4 may lead to conflated effect, we did not include them in the interaction analysis.

### Genotype data processing

We used the imputed genotype dataset released by UK Biobank in 2017.^19^ About 92 million autosomal variants were imputed with the Haplotype Reference Consortium (HRC) panel and the UK10K + 1000 Genomes panel as references. Participants were removed if they had mismatches between self-reported sex and genetically inferred sex, exhibited sex chromosome aneuploidy, or were identified as outliers for heterozygosity or missing rate. We also excluded participants who had excessive third-degree relatives and randomly removed one individual among pairs of related individuals closer than the third degree.

For marker-based quality control, we first filtered out low-quality imputed variants with an info score below 0.3 and retained only biallelic variants. SNPs with a minor allele frequency (MAF) less than 0.1%, a missing calling rate larger than 5%, and a Hardy-Weinberg equilibrium P-value less than 10^-8^ were also removed. Finally, we ensured that the missing call rate per individual was below 2% after these processes. Genotype data was all processed by PLINK version 1.9 or version 2.0.^20–22^

### Candidate SNPs selection

We performed time-to-event GWAS by a scalable and accurate method, saddlepoint approximation implementation based on the Cox proportional hazards regression model (SPACox), for all-cause dementia, Alzheimer’s disease, and vascular dementia, respectively.^17^ Age, sex, and the top 10 genetic PCs were included as covariates. Independent loci of genome-wide significant signals (p-value < 5e-8) for each outcome were defined based on clumping (p-value < 5e-8 and r² < 0.1 within a 250 kb physical distance) and then merging if two signal clusters are within 250 kb of each other. The novelty of these loci was defined with comparison to previous GWAS on the development of ADRD.^4^

Since we are looking for genetic factors that modify the association of fish oil supplementation and dementia outcomes, the associations of these genetic factors with the dementia outcomes are dependent on the fish oil supplementation status and may be masked or weakened in the GWAS in the entire dataset. Therefore, we also performed time-to-event GWAS of each outcome in the two subgroups of fish oil users and non-users, separately. Candidate variants for subsequent interaction analysis were selected using a lenient threshold of p-value < 1e-5 from GWAS in the entire dataset or in the two subgroups.

### Time-to-event G×E interaction analysis

Interactions between candidate SNPs and fish oil supplementation on incident dementia were evaluated using the Cox proportional hazards models. Each model was adjusted by age, sex, the top 10 genetic PCs, fish oil supplementation, and the corresponding SNP. Interaction signals were identified with a Bonferroni-corrected threshold based on the number of independent candidate loci. These loci were obtained from the clumped regions of suggestive signals (r² < 0.1, p-value < 1e-5 within a 250 kb physical distance) with ±250 kb extension. The interaction analyses with *APOE* ε4 dosage were performed as positive controls. Furthermore, considering the potential effects of lifestyle, dietary patterns, and related medical history, we performed multiple sensitivity analyses by adjusting for additional covariates. Details were described in the Supplementary Methods.

### Gene set enrichment analysis

Genome-wide significant interaction signals were annotated by the Variant Effect Predictor (VEP) toolset on the Ensembl platform.^23^ Each candidate variant was assigned location information for the genes, transcripts, and protein sequences that it might affect. Then, gene set enrichment analysis was conducted on the Web-based Cell-type-Specific Enrichment Analysis of Genes (WebCSEA) website for the list of candidate protein-coding genes included in the WebCSEA background gene list, to identify the tissue or cell-type-specific expression signatures of these interaction signals.^24^

### Statistical analysis

Descriptive and inferential analyses were performed using R version 3.4.1.^25^ Imputation of missing covariate data was conducted by the *mice* package using predictive mean matching, logistic regression, and polytomous logistic regression methods, respectively, for continuous variables, binary variables, and unordered multiple categorial variables.^26^

## Results

As shown in **Figure 1**, 425,157 participants from the UK Biobank are of genetic European ancestry, completed the touchscreen questionnaire on fish oil supplementation, and did not have a diagnosis of dementia at recruitment. After quality control with genotype data and removing related individuals, 357,631 participants with a mean age of 56.81±8.01 were included in our study. Around 31.7% of participants reported consuming fish oil supplements, as documented in the touchscreen questionnaire at baseline (**Table S1-S2**). Older individuals and women appeared more likely to take fish oil supplements.

**Figure 1.**
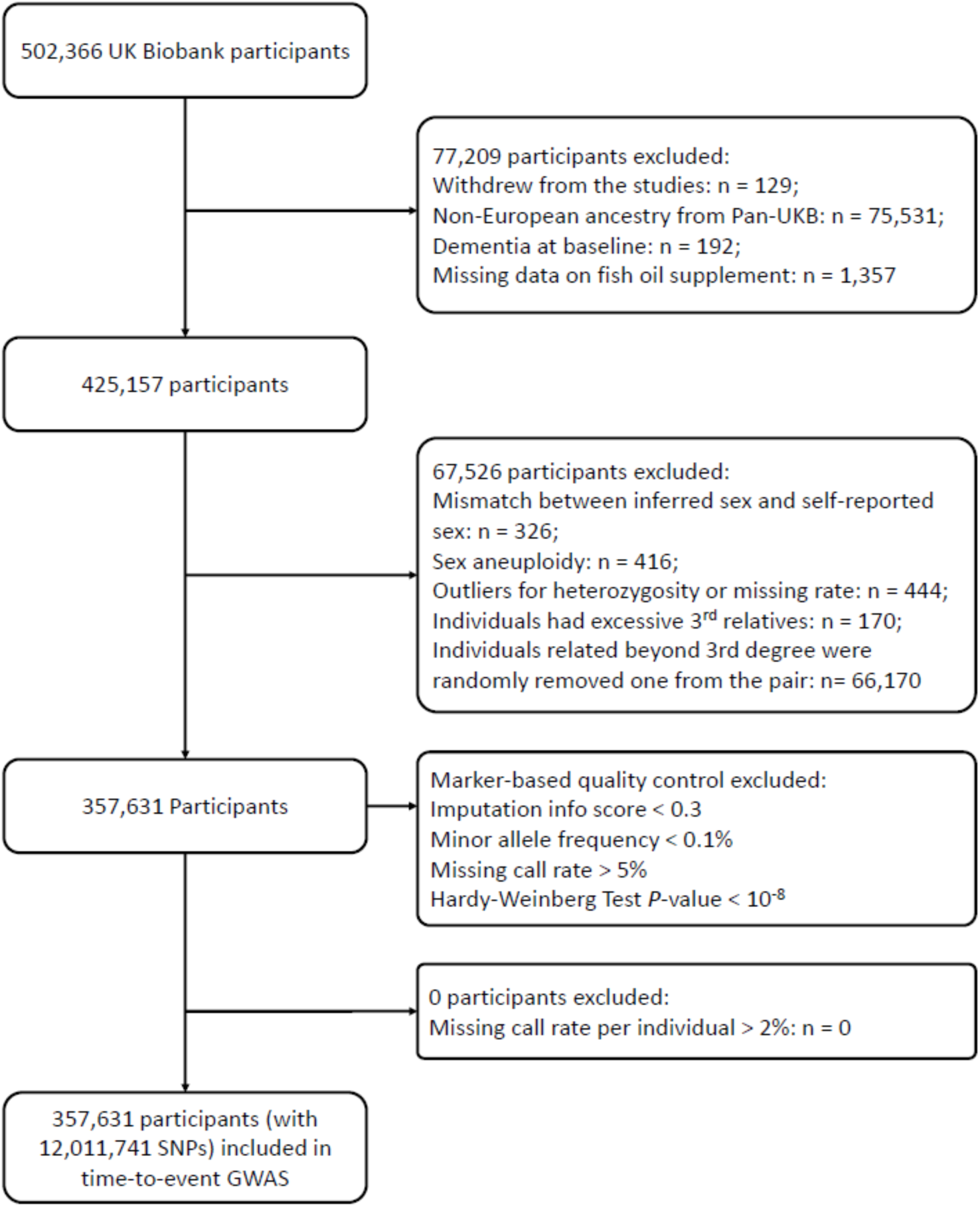
Flow chart of participant inclusion and exclusion

### Associations between fish oil supplementation and incident dementias

We evaluated the associations between fish oil supplementation and incident all-cause dementia and its subtypes using Cox proportional hazards models over a median follow-up period of 13.8 years. Regular intake of fish oil supplements demonstrated protective associations with incident all-cause dementia (HR, 0.92; 95 % CI, 0.87–0.96) and vascular dementia (HR, 0.80; 95 % CI, 0.72–0.89) after adjustment for age and sex (**Table S3**). However, fish oil supplementation showed no significant association with the onset of Alzheimer’s disease.

### Time-to-event GWAS on dementias

We first performed time-to-event GWAS on three dementia outcomes separately, to identify disease-related loci and select candidate variants for further interaction analysis (**Figure 2A**). In the entire dataset of 357,631 participants and at the genome-wide significance (p-value <5e-8), we identified 6 loci for all-cause dementia, 5 for Alzheimer’s disease, and 2 for vascular dementia (**Table S4**). *APOE* was the locus shared among these three dementia outcomes. Bridging integrator 1 (*BIN1*), triggering receptor expressed on myeloid cells 2 (*TREM2*), and zinc finger CW-type (*ZCWPW1*)/PWWP domain containing 1 (*NYAP1*) were identified in both all-cause dementia and Alzheimer’s disease outcomes. Protein tyrosine kinase 2 beta (*PTK2B*)/clusterin (*CLU*) and membrane-spanning 4A (*MS4A*) were significantly associated with the onset of all-cause dementia, while aph-1 homolog B (*APH1B*) was specifically linked to Alzheimer’s disease. A novel locus for vascular dementia was discovered on chromosome 1, with castor zinc finger 1 (*CASZ1*) being the closest gene to the top signal.

**Figure 2.**
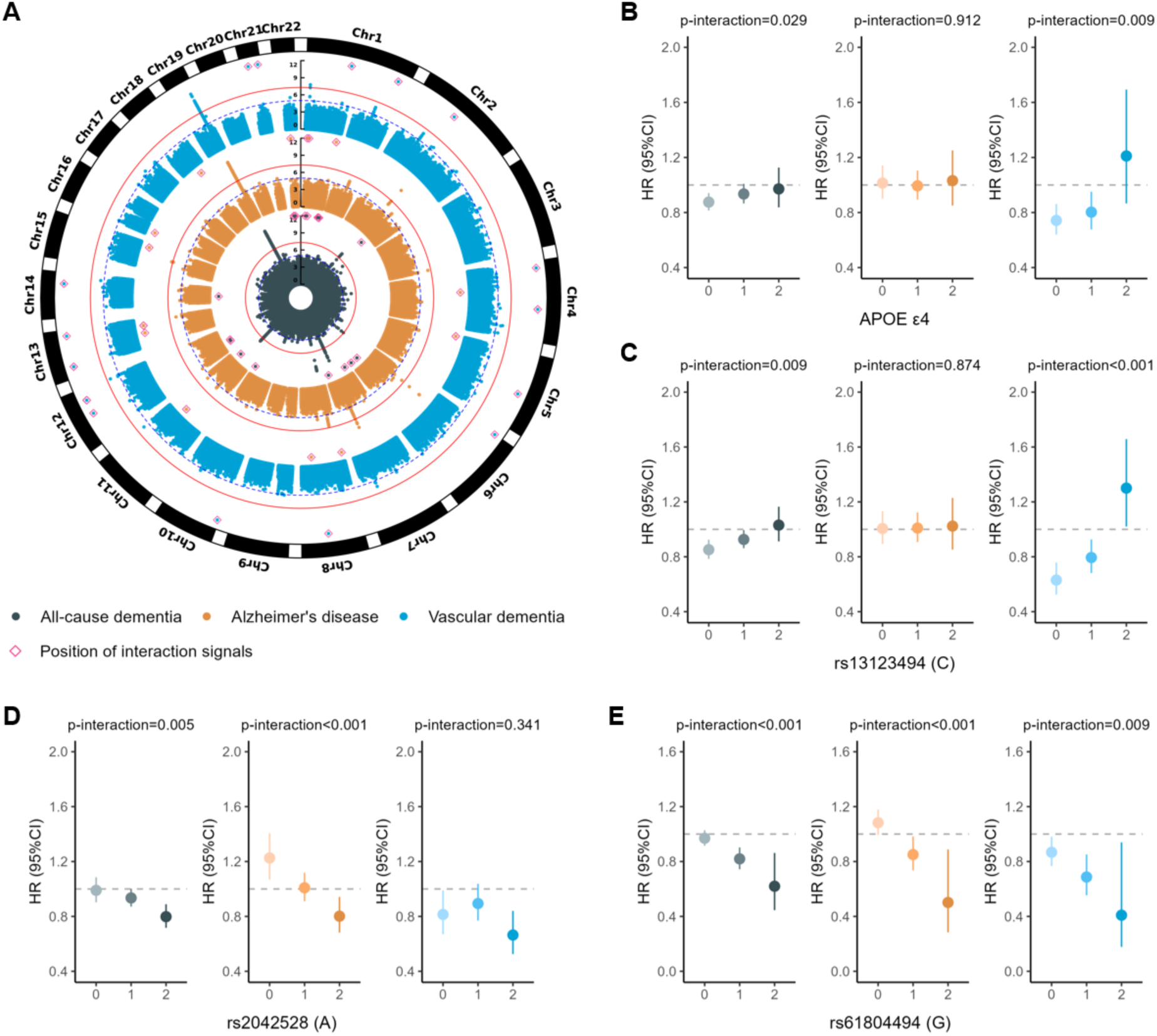
Genome-wide association and G × fish oil supplementation interaction analysis of dementias. A) Manhattan plots of time-to-event GWAS for all-cause dementia (dark cyan), Alzheimer’s disease (orange), and vascular dementia (blue). Variants with a -log10(p-value) below 11 are not shown. Thresholds of genome-wide significance (P-value< 5e-8) and suggestive significance (P-value< 1e-5) were indicated by solid red lines and dashed blue lines, respectively. Positions of interaction signals were labeled on the top of Manhattan plots. B) - E) Associations between regular fish oil supplementation and incident dementias in *APOE* ε4 genotypes (B), rs13123494 genotypes (C), rs2042528 genotypes (D), and rs61804494 genotypes (E). The grey dashed lines represented the lines of null effect. The values of p-interaction were evaluated for the interaction terms SNP×fish oil supplementation (Yes/No) by Cox regression models adjusted by age, sex, and top 10 genetic PCs.

Considering that genetic variants with interaction effects have exposure-dependent effects on the onset of dementia, we conducted stratified GWAS for the two subgroups of fish oil users and non-users. A total of 1,819 suggestive common SNPs (p-value<1e-5; MAF > 0.01) in 178 independent loci were then selected for interaction analysis (**Figure S1-S3**).

### Time-to-event Gene × Fish Oil interaction analysis

We performed time-to-event gene × fish oil interaction analysis with the selected candidate SNPs for all-cause dementia, Alzheimer’s disease, and vascular dementia. Our primary interaction analyses used the Cox proportional hazards model, referred to as Model 1, which adjusted for age, sex, status of fish oil supplementation, and dosage of the corresponding SNP. A total of 81 SNPs in 43 loci were found to have significant interaction signals (p-value<2.8e-4, or 0.05/178 candidate loci) for one of the three dementia outcomes (**Figure 2A**, **Table 1**, **Table S5-S8**). A total of 22 protein-coding genes from these interaction loci were annotated through the VEP tool. One locus was annotated to the calcium/calmodulin-dependent protein kinase II delta (*CAMK2D*) gene, which is highly expressed in heart muscle tissue. The top variant at this locus (rs13123494) exhibited a pattern similar to that of *APOE* ε4, but with stronger effect sizes and statistical significance (**Figure 2B** and **2C**). Among individuals with no or one C allele, taking fish oil supplements was associated with lower risk of all-cause dementia and vascular dementia, whereas a risk-increasing association was observed in those homozygous for the C allele. The association between AD and fish oil supplementation did not vary across different genotypes. Other interaction loci, such as rs2042528 and rs61804494, with different interaction patterns than that of *APOE* ε4, were also shown in **Figure 2D** and **2E**.

**Table 1.**
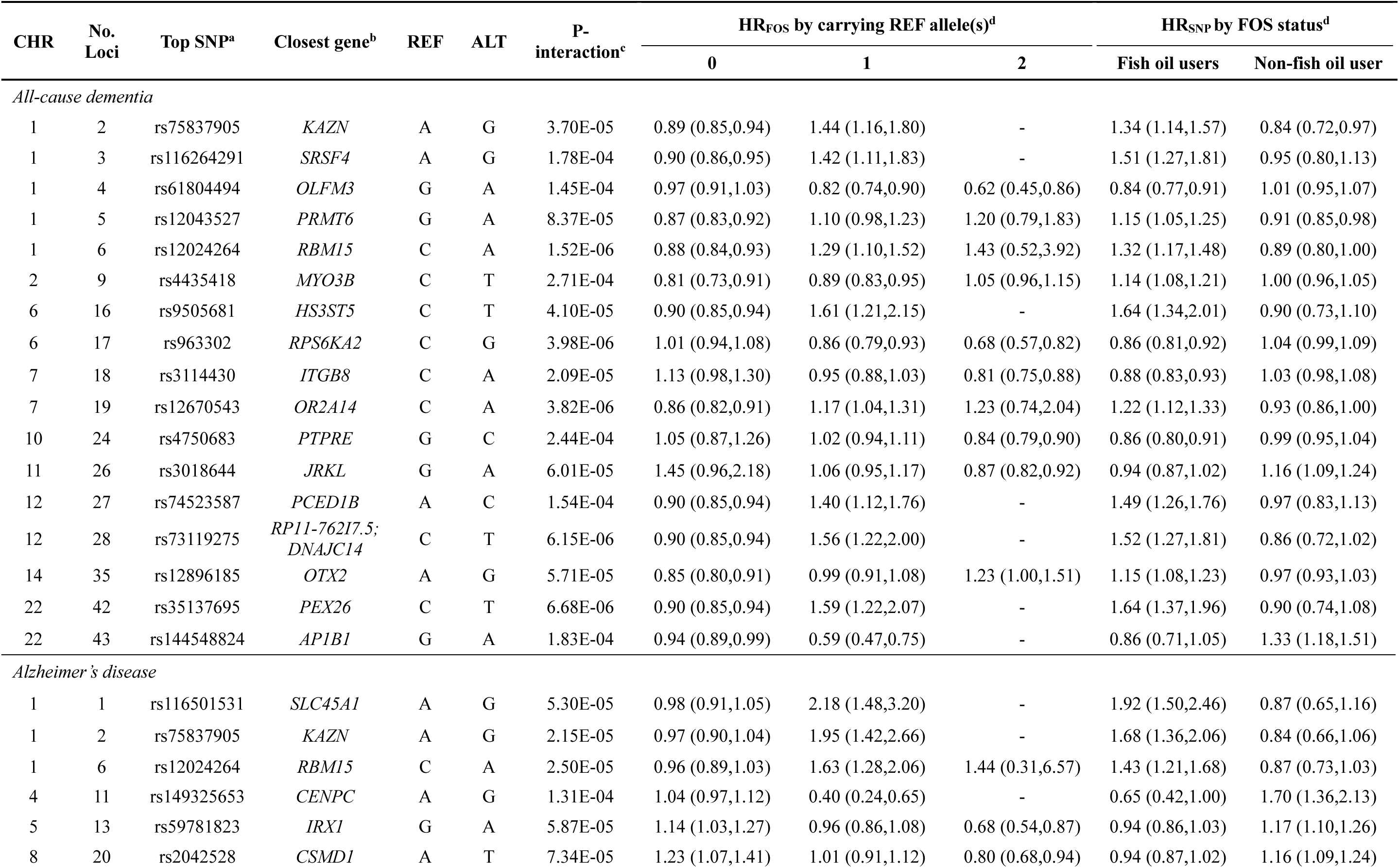
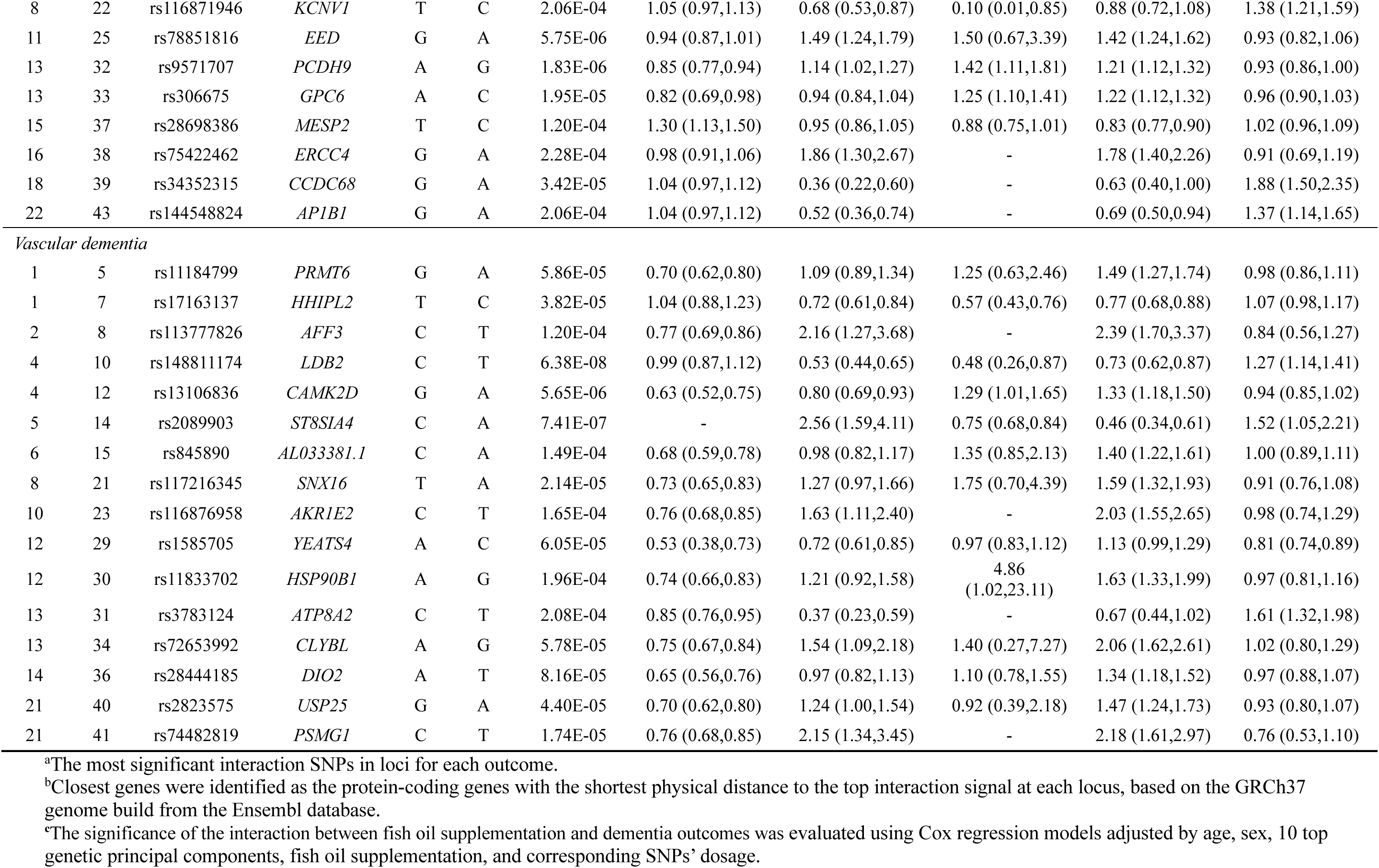
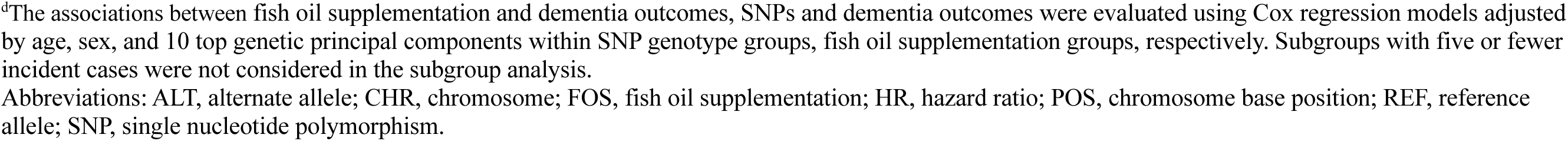
Genetic loci with significant gene-fish oil interaction effects on three dementia outcomes.

| CHR | No. Loci | Top SNP<sup>a</sup> | Closest gene<sup>b</sup> | REF | ALT | P-interaction<sup>c</sup> | HR<sub>FOS</sub> by carrying REF allele(s)<sup>d</sup> |  |  | HR<sub>SNP</sub> by FOS status<sup>d</sup> |  |
| --- | --- | --- | --- | --- | --- | --- | --- | --- | --- | --- | --- |
|  |  |  |  |  |  |  | 0 | 1 | 2 | Fish oil users | Non-fish oil user |
| All-cause dementia |  |  |  |  |  |  |  |  |  |  |  |
| 1 | 2 | rs75837905 | KAZN | A | G | 3.70E-05 | 0.89 (0.85,0.94) | 1.44 (1.16,1.80) | - | 1.34 (1.14,1.57) | 0.84 (0.72,0.97) |
| 1 | 3 | rs116264291 | SRSF4 | A | G | 1.78E-04 | 0.90 (0.86,0.95) | 1.42 (1.11,1.83) | - | 1.51 (1.27,1.81) | 0.95 (0.80,1.13) |
| 1 | 4 | rs61804494 | OLFM3 | G | A | 1.45E-04 | 0.97 (0.91,1.03) | 0.82 (0.74,0.90) | 0.62 (0.45,0.86) | 0.84 (0.77,0.91) | 1.01 (0.95,1.07) |
| 1 | 5 | rs12043527 | PRMT6 | G | A | 8.37E-05 | 0.87 (0.83,0.92) | 1.10 (0.98,1.23) | 1.20 (0.79,1.83) | 1.15 (1.05,1.25) | 0.91 (0.85,0.98) |
| 1 | 6 | rs12024264 | RBM15 | C | A | 1.52E-06 | 0.88 (0.84,0.93) | 1.29 (1.10,1.52) | 1.43 (0.52,3.92) | 1.32 (1.17,1.48) | 0.89 (0.80,1.00) |
| 2 | 9 | rs4435418 | MYO3B | C | T | 2.71E-04 | 0.81 (0.73,0.91) | 0.89 (0.83,0.95) | 1.05 (0.96,1.15) | 1.14 (1.08,1.21) | 1.00 (0.96,1.05) |
| 6 | 16 | rs9505681 | HS3ST5 | C | T | 4.10E-05 | 0.90 (0.85,0.94) | 1.61 (1.21,2.15) | - | 1.64 (1.34,2.01) | 0.90 (0.73,1.10) |
| 6 | 17 | rs963302 | RPS6KA2 | C | G | 3.98E-06 | 1.01 (0.94,1.08) | 0.86 (0.79,0.93) | 0.68 (0.57,0.82) | 0.86 (0.81,0.92) | 1.04 (0.99,1.09) |
| 7 | 18 | rs3114430 | ITGB8 | C | A | 2.09E-05 | 1.13 (0.98,1.30) | 0.95 (0.88,1.03) | 0.81 (0.75,0.88) | 0.88 (0.83,0.93) | 1.03 (0.98,1.08) |
| 7 | 19 | rs12670543 | OR2A14 | C | A | 3.82E-06 | 0.86 (0.82,0.91) | 1.17 (1.04,1.31) | 1.23 (0.74,2.04) | 1.22 (1.12,1.33) | 0.93 (0.86,1.00) |
| 10 | 24 | rs4750683 | PTPRE | G | C | 2.44E-04 | 1.05 (0.87,1.26) | 1.02 (0.94,1.11) | 0.84 (0.79,0.90) | 0.86 (0.80,0.91) | 0.99 (0.95,1.04) |
| 11 | 26 | rs3018644 | JRKL | G | A | 6.01E-05 | 1.45 (0.96,2.18) | 1.06 (0.95,1.17) | 0.87 (0.82,0.92) | 0.94 (0.87,1.02) | 1.16 (1.09,1.24) |
| 12 | 27 | rs74523587 | PCED1B | A | C | 1.54E-04 | 0.90 (0.85,0.94) | 1.40 (1.12,1.76) | - | 1.49 (1.26,1.76) | 0.97 (0.83,1.13) |
| 12 | 28 | rs73119275 | RP11-762I7.5;<br>DNAJC14 | C | T | 6.15E-06 | 0.90 (0.85,0.94) | 1.56 (1.22,2.00) | - | 1.52 (1.27,1.81) | 0.86 (0.72,1.02) |
| 14 | 35 | rs12896185 | OTX2 | A | G | 5.71E-05 | 0.85 (0.80,0.91) | 0.99 (0.91,1.08) | 1.23 (1.00,1.51) | 1.15 (1.08,1.23) | 0.97 (0.93,1.03) |
| 22 | 42 | rs35137695 | PEX26 | C | T | 6.68E-06 | 0.90 (0.85,0.94) | 1.59 (1.22,2.07) | - | 1.64 (1.37,1.96) | 0.90 (0.74,1.08) |
| 22 | 43 | rs144548824 | AP1B1 | G | A | 1.83E-04 | 0.94 (0.89,0.99) | 0.59 (0.47,0.75) | - | 0.86 (0.71,1.05) | 1.33 (1.18,1.51) |
| Alzheimer's disease |  |  |  |  |  |  |  |  |  |  |  |
| 1 | 1 | rs116501531 | SLC45A1 | A | G | 5.30E-05 | 0.98 (0.91,1.05) | 2.18 (1.48,3.20) | - | 1.92 (1.50,2.46) | 0.87 (0.65,1.16) |
| 1 | 2 | rs75837905 | KAZN | A | G | 2.15E-05 | 0.97 (0.90,1.04) | 1.95 (1.42,2.66) | - | 1.68 (1.36,2.06) | 0.84 (0.66,1.06) |
| 1 | 6 | rs12024264 | RBM15 | C | A | 2.50E-05 | 0.96 (0.89,1.03) | 1.63 (1.28,2.06) | 1.44 (0.31,6.57) | 1.43 (1.21,1.68) | 0.87 (0.73,1.03) |
| 4 | 11 | rs149325653 | CENPC | A | G | 1.31E-04 | 1.04 (0.97,1.12) | 0.40 (0.24,0.65) | - | 0.65 (0.42,1.00) | 1.70 (1.36,2.13) |
| 5 | 13 | rs59781823 | IRX1 | G | A | 5.87E-05 | 1.14 (1.03,1.27) | 0.96 (0.86,1.08) | 0.68 (0.54,0.87) | 0.94 (0.86,1.03) | 1.17 (1.10,1.26) |
| 8 | 20 | rs2042528 | CSMD1 | A | T | 7.34E-05 | 1.23 (1.07,1.41) | 1.01 (0.91,1.12) | 0.80 (0.68,0.94) | 0.94 (0.87,1.02) | 1.16 (1.09,1.24) |
| 8 | 22 | rs116871946 | *KCNV1* | T | C | 2.06E-04 | 1.05 (0.97,1.13) | 0.68 (0.53,0.87) | 0.10 (0.01,0.85) | 0.88 (0.72,1.08) | 1.38 (1.21,1.59) |
| 11 | 25 | rs78851816 | *EED* | G | A | 5.75E-06 | 0.94 (0.87,1.01) | 1.49 (1.24,1.79) | 1.50 (0.67,3.39) | 1.42 (1.24,1.62) | 0.93 (0.82,1.06) |
| 13 | 32 | rs9571707 | *PCDH9* | A | G | 1.83E-06 | 0.85 (0.77,0.94) | 1.14 (1.02,1.27) | 1.42 (1.11,1.81) | 1.21 (1.12,1.32) | 0.93 (0.86,1.00) |
| 13 | 33 | rs306675 | *GPC6* | A | C | 1.95E-05 | 0.82 (0.69,0.98) | 0.94 (0.84,1.04) | 1.25 (1.10,1.41) | 1.22 (1.12,1.32) | 0.96 (0.90,1.03) |
| 15 | 37 | rs28698386 | *MESP2* | T | C | 1.20E-04 | 1.30 (1.13,1.50) | 0.95 (0.86,1.05) | 0.88 (0.75,1.01) | 0.83 (0.77,0.90) | 1.02 (0.96,1.09) |
| 16 | 38 | rs75422462 | *ERCC4* | G | A | 2.28E-04 | 0.98 (0.91,1.06) | 1.86 (1.30,2.67) | - | 1.78 (1.40,2.26) | 0.91 (0.69,1.19) |
| 18 | 39 | rs34352315 | *CCDC68* | G | A | 3.42E-05 | 1.04 (0.97,1.12) | 0.36 (0.22,0.60) | - | 0.63 (0.40,1.00) | 1.88 (1.50,2.35) |
| 22 | 43 | rs144548824 | *AP1B1* | G | A | 2.06E-04 | 1.04 (0.97,1.12) | 0.52 (0.36,0.74) | - | 0.69 (0.50,0.94) | 1.37 (1.14,1.65) |
| *Vascular dementia* |  |  |  |  |  |  |  |  |  |  |  |
| 1 | 5 | rs11184799 | *PRMT6* | G | A | 5.86E-05 | 0.70 (0.62,0.80) | 1.09 (0.89,1.34) | 1.25 (0.63,2.46) | 1.49 (1.27,1.74) | 0.98 (0.86,1.11) |
| 1 | 7 | rs17163137 | *HHIPL2* | T | C | 3.82E-05 | 1.04 (0.88,1.23) | 0.72 (0.61,0.84) | 0.57 (0.43,0.76) | 0.77 (0.68,0.88) | 1.07 (0.98,1.17) |
| 2 | 8 | rs113777826 | *AFF3* | C | T | 1.20E-04 | 0.77 (0.69,0.86) | 2.16 (1.27,3.68) | - | 2.39 (1.70,3.37) | 0.84 (0.56,1.27) |
| 4 | 10 | rs148811174 | *LDB2* | C | T | 6.38E-08 | 0.99 (0.87,1.12) | 0.53 (0.44,0.65) | 0.48 (0.26,0.87) | 0.73 (0.62,0.87) | 1.27 (1.14,1.41) |
| 4 | 12 | rs13106836 | *CAMK2D* | G | A | 5.65E-06 | 0.63 (0.52,0.75) | 0.80 (0.69,0.93) | 1.29 (1.01,1.65) | 1.33 (1.18,1.50) | 0.94 (0.85,1.02) |
| 5 | 14 | rs2089903 | *ST8SIA4* | C | A | 7.41E-07 | - | 2.56 (1.59,4.11) | 0.75 (0.68,0.84) | 0.46 (0.34,0.61) | 1.52 (1.05,2.21) |
| 6 | 15 | rs845890 | *AL033381.1* | C | A | 1.49E-04 | 0.68 (0.59,0.78) | 0.98 (0.82,1.17) | 1.35 (0.85,2.13) | 1.40 (1.22,1.61) | 1.00 (0.89,1.11) |
| 8 | 21 | rs117216345 | *SNX16* | T | A | 2.14E-05 | 0.73 (0.65,0.83) | 1.27 (0.97,1.66) | 1.75 (0.70,4.39) | 1.59 (1.32,1.93) | 0.91 (0.76,1.08) |
| 10 | 23 | rs116876958 | *AKR1E2* | C | T | 1.65E-04 | 0.76 (0.68,0.85) | 1.63 (1.11,2.40) | - | 2.03 (1.55,2.65) | 0.98 (0.74,1.29) |
| 12 | 29 | rs1585705 | *YEATS4* | A | C | 6.05E-05 | 0.53 (0.38,0.73) | 0.72 (0.61,0.85) | 0.97 (0.83,1.12) | 1.13 (0.99,1.29) | 0.81 (0.74,0.89) |
| 12 | 30 | rs11833702 | *HSP90B1* | A | G | 1.96E-04 | 0.74 (0.66,0.83) | 1.21 (0.92,1.58) | 4.86<br>(1.02,23.11) | 1.63 (1.33,1.99) | 0.97 (0.81,1.16) |
| 13 | 31 | rs3783124 | *ATP8A2* | C | T | 2.08E-04 | 0.85 (0.76,0.95) | 0.37 (0.23,0.59) | - | 0.67 (0.44,1.02) | 1.61 (1.32,1.98) |
| 13 | 34 | rs72653992 | *CLYBL* | A | G | 5.78E-05 | 0.75 (0.67,0.84) | 1.54 (1.09,2.18) | 1.40 (0.27,7.27) | 2.06 (1.62,2.61) | 1.02 (0.80,1.29) |
| 14 | 36 | rs28444185 | *DIO2* | A | T | 8.16E-05 | 0.65 (0.56,0.76) | 0.97 (0.82,1.13) | 1.10 (0.78,1.55) | 1.34 (1.18,1.52) | 0.97 (0.88,1.07) |
| 21 | 40 | rs2823575 | *USP25* | G | A | 4.40E-05 | 0.70 (0.62,0.80) | 1.24 (1.00,1.54) | 0.92 (0.39,2.18) | 1.47 (1.24,1.73) | 0.93 (0.80,1.07) |
| 21 | 41 | rs74482819 | *PSMG1* | C | T | 1.74E-05 | 0.76 (0.68,0.85) | 2.15 (1.34,3.45) | - | 2.18 (1.61,2.97) | 0.76 (0.53,1.10) |

<sup>a</sup>The most significant interaction SNPs in loci for each outcome.
<sup>b</sup>Closest genes were identified as the protein-coding genes with the shortest physical distance to the top interaction signal at each locus, based on the GRCh37 genome build from the Ensembl database.
<sup>c</sup>The significance of the interaction between fish oil supplementation and dementia outcomes was evaluated using Cox regression models adjusted by age, sex, 10 top genetic principal components, fish oil supplementation, and corresponding SNPs' dosage.
<sup>d</sup>The associations between fish oil supplementation and dementia outcomes, SNPs and dementia outcomes were evaluated using Cox regression models adjusted by age, sex, and 10 top genetic principal components within SNP genotype groups, fish oil supplementation groups, respectively. Subgroups with five or fewer incident cases were not considered in the subgroup analysis.
Abbreviations: ALT, alternate allele; CHR, chromosome; FOS, fish oil supplementation; HR, hazard ratio; POS, chromosome base position; REF, reference allele; SNP, single nucleotide polymorphism.

We compared the GWAS loci and interaction loci. One locus on chromosome 11 overlapped with the reported ADRD locus embryonic ectoderm development (*EED*), and two loci on chromosomes 1 and 21 overlapped with known GWAS loci, serine and arginine rich splicing factor 4 (*SRSF4*) and proteasome assembly chaperone 1 (*PSMG1*), for polyunsaturated fatty acids (PUFAs) (**Table S8**). Of note, none of the interaction loci overlapped with the genome-wide significant loci in our time-to-event GWAS on dementias (**Table S9**).

While the total number of interaction loci across the three dementia outcomes was 43, the numbers were 17, 14, and 16, respectively, for all-cause dementia, Alzheimer’s disease, and vascular dementia. Among them, 15, 12, and 15 loci reached suggestive significance in GWAS in either fish oil subgroup. Interestingly, these interaction loci demonstrated a fish oil-specific genetic association pattern. 13 out of the 15 loci (87%, p-value = 0.0074) for all-cause dementia and 12 out of the 15 loci for vascular dementia (80%, p-value = 0.035) showing suggestive significance exclusively in the fish oil subgroup GWAS but not in the non-fish oil users. Alzheimer’s disease did not display such a preference, with 7 out of 12 loci (58%) exhibiting suggestive significance only in fish oil users (**Table S9, Figure S4**).

### Sensitivity analyses

We performed a series of sensitivity analyses to evaluate the robustness of the identified interaction signals by adjusting for socioeconomic status, lifestyle, dietary patterns, and related medical history. Models adjusting for some or all of these additional covariates yielded similarly significant interaction signals (**Figure S5**). We also compared the interaction signals from Model 1 by using different questionnaires to define fish oil supplementation (**Figure S6**). Interaction signals on peroxisomal biogenesis factor 26 (*PEX26*) locus, *EED* locus, and *CAMK2D* locus were replicated in all-cause dementia, AD, and vascular dementia outcomes, respectively.

### Gene set enrichment analysis

Gene set enrichment analysis was performed on 21 protein-coding genes included in the background gene list, to identify their expression signatures across 111 human tissue panels and 1,355 tissue-cell types spanning 12 human organ systems using the WebCSEA website. These 21 genes were highly expressed in various cell types of the nervous system (**Figure 3**). Notably, they were enriched in layer 6b excitatory neuron subtypes (Ex6b) in the frontal and visual cortex (p-value < 1.67e-4 and 4.64e-4, respectively), oligodendrocyte progenitor cells (OPCs) in the cerebellar hemisphere of adults (p-value < 5.39e-4), and oligodendrocytes in both the cerebrum and cerebellum (p-value < 1.49e-4 and 1.01e-3, respectively), as well as astrocytes in the cerebellum of fetuses (p-value < 2.56e-3) through permutation-based tests (**Figure S7**).

**Figure 3.**
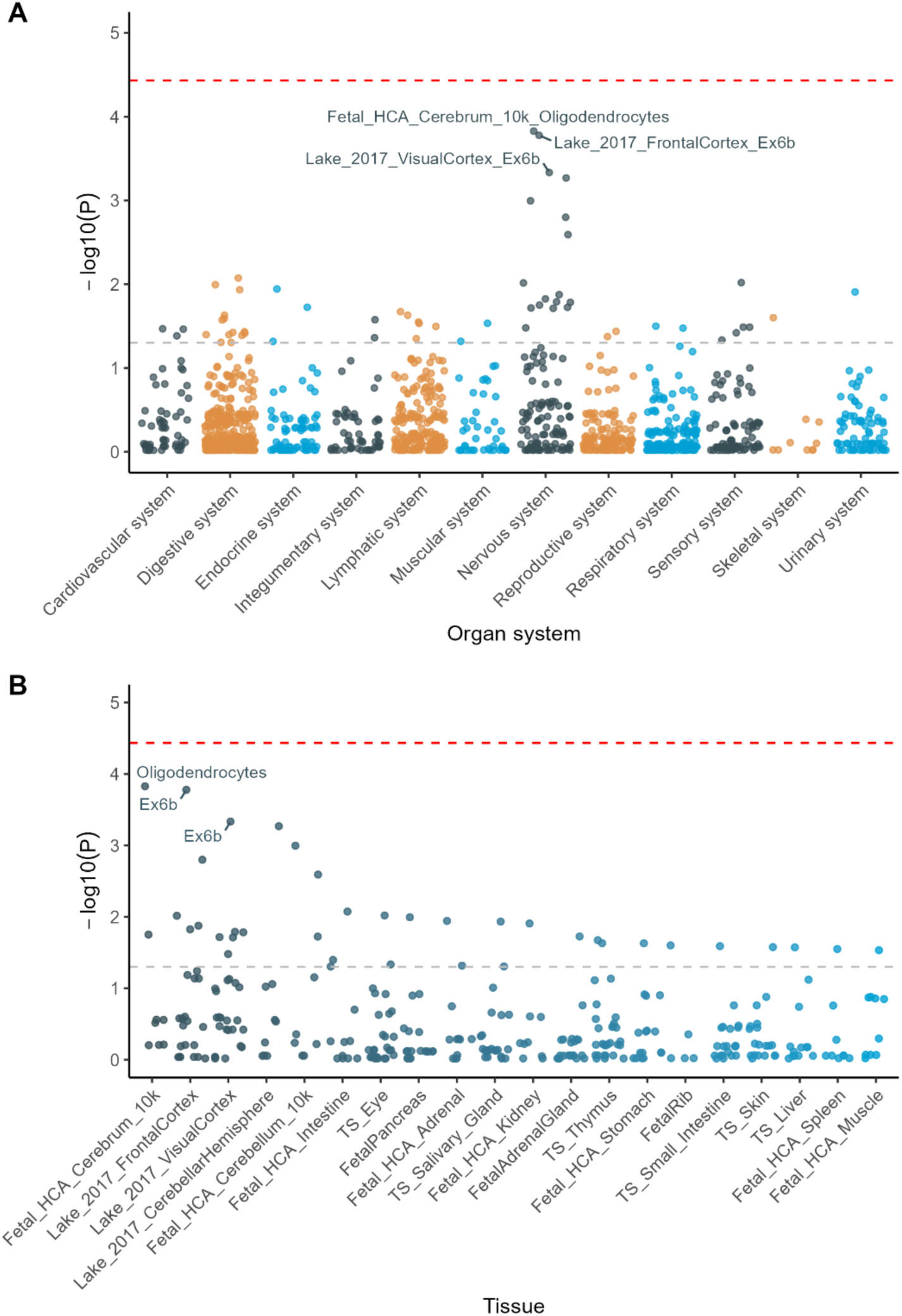
Protein-coding genes enrichment in tissues or cell types. A) Tissue-cell types of genes specifically expressed across overall organ systems. B) Top 20 tissues with specific general cell types where the target genes were highly enriched. P-values of cell-type-specific enrichment analysis after normalizing the effect of gene set length by permutation-based method. Red and grey dashed lines represent thresholds for Bonferroni-corrected significance (0.05/total number of tissue-cell types) and nominal significance (0.05), separately.

## Discussion

In this study, we identified 6, 5, and 2 independent loci, respectively, associated with incident all-cause dementia, Alzheimer’s disease, and vascular dementia. Most of these loci were reported in the previous ADRD GWAS meta-analysis.^4^ Among 178 suggestive GWAS loci, 43 interaction signals significantly modified the association between fish oil supplementation and dementia development. Three loci were previously known to be associated with ADRD or PUFAs.^4,27–31^ However, none of these loci overlapped with the time-to-event GWAS loci we found, indicating that the interaction loci may have been missed in typical GWAS. Additionally, candidate protein-coding genes at interaction loci are highly expressed in the nervous system, especially in some specific neuron tissues and cell types. These results support that the new interaction signals may be involved in the pathway of brain functioning and PUFA metabolism, contributing to dementia development.

*APOE* were the strongest loci related to all-cause dementia and its two major subtypes in the time-to-event GWAS. It plays a key role in lipid metabolism and homeostasis. One of the major isoforms, *APOE* ε4 was known to be positively associated with amyloid plaques and neurofibrillary tangles (NFTs) formation, contributing to AD pathogenesis.^32,33^ Other known AD associated genes, *BIN1*, *TREM2*, *ZCWPW1*, and *APH1B*, were also replicated in our study.^4^ These genes were differentially expressed in specific cell types in the brains of AD patients compared to healthy individuals.^34^ Additionally, we identified a novel locus exclusively associated with vascular dementia. The top SNP within this locus is an intergenic variant likely involved in gene regulation. Castor zinc finger 1 (*CASZ1*), located in this region, plays roles in both neural and cardiovascular development.^35^ Other genes at this unknown locus, such as TAR DNA binding protein (*TARDBP*) and MBL associated serine protease 2 (*MASP2*), showed significant interactions with genes involved in tau-related pathologies.^36^

We identified 43 interaction loci modifying the associations between fish oil supplementation and dementia at statistical significance stronger than that of *APOE* ε4. These findings suggested a need and potential strategies for personalizing fish oil recommendations for high-risk populations. Among the interaction loci, *EED*, *SRSF4*, and *PSMG1*, were reported to be associated with incident ADD and PUFA traits, omega-3 PUFA level or percentage, and the ratio of omega-6 to omega-3 PUFA levels. These suggest that the interaction signals may be involved in dementia pathology and PUFA metabolism, although the specific mechanism is still unclear. Notably, none of the interaction loci overlapped with the time-to-event GWAS loci mentioned earlier. This disparity indicated that directly investigating G×E relationships based solely on GWAS hits may have missed these signals. Instead, we included candidate SNPs from subgroup-specific GWAS to consider the possibility of different or opposite effects of SNPs on outcomes between fish oil users and non-users, increasing the chances of catching the interaction signals. Previous interaction analyses between fish oil and polygenic risk score (PRS) of AD, excluding the effect of *APOE ε4*, showed no significant results in all-cause dementia and AD development.^15^ These also indicate the differences in the effects of GWAS loci and interaction loci in dementia and call for the consideration of G×E when applying PRS for disease prediction.

Protein-coding genes affected by interaction signals were enriched in high expression in oligodendrocytes, OPCs, and Ex6b cell types in adult brain tissues according to the gene set enrichment analysis. OPCs are known as oligodendrocyte precursor cells, which would divide and regenerate oligodendrocytes for myelin repair and myelination. This process slows down with aging and is associated with reduced myelination in AD, contributing to increased cognitive decline.^37–39^ The balance of excitatory and inhibitory inputs onto cortical neurons is strictly regulated by homeostatic mechanisms.^40^ A recent study revealed that the positive feedback loop between hyperexcitability and amyloidosis may lead to neuronal dysfunction and neurodegeneration.^41^ Additionally, omega-3 fatty acids have also been reported to protect against myelin sheath damage and affect synaptic plasticity.^42,43^ Therefore, we speculated that genetic signaling and fish oil supplementation may play some synergistic roles in myelin maintenance and synaptic homeostasis. Furthermore, we found that half of the interaction signals were mainly located in intronic or intergenic regions, suggesting gene regulation as their possible molecular mechanisms.

In this study, we mainly focused on all-cause dementia and its two subtypes, AD and vascular dementia, in the European population due to the limitation of sample and case sizes. Further studies on different populations and dementia subtypes, such as frontotemporal dementia, are needed to explore similarities and differences in dementia pathogenesis and across ancestral groups. Moreover, we applied a two-step approach to select a subset of SNPs for further time-to-event interaction analysis, reducing the computational cost of genome-wide interaction analysis of longitudinal outcomes. However, this approach was limited to categorial exposure and had the possibility of missing potential interaction signals during the candidate loci selection process. More effective algorithms need to be developed for broader and faster screening. Additionally, the binary fish oil exposure lacks information about dose or supplementation duration. Likely due to this imperfect exposure measurement, our results from sensitivity analyses with fish oil supplementation from different questionnaires only had limited replication. More reliable indicators of fish oil supplementation or dietary omega-3 intake, such as the circulating omega-3 fatty acid level, may be used for future studies as validation.

## Conclusions

We identified 43 genomic loci that modify the association between fish oil supplementation and dementias. Gene set enrichment analysis showed that protein-coding genes around the interaction signals demonstrated high expression signatures in several neuron cell types in brain tissues. This study revealed potential interaction relationships between genetic factors and fish oil supplementation on the development of dementias. It provides new insight into the roles of fatty acids in disease pathogenesis and calls for genome-informed personalized dietary recommendations for dementia prevention.

## Data Availability

The datasets analyzed during the current study are available from the UK Biobank through an application process.

https://www.ukbiobank.ac.uk/

## Acknowledgements

This research has been conducted using the UK Biobank Resource under Application Number 48818. This work uses data provided by patients and collected by the NHS as part of their care and support. These data are copyrighted, 2022, NHS England. Reused with the permission of the NHS England and UK Biobank. All rights reserved. This research used data assets made available by National Safe Haven as part of the Data and Connectivity National Core Study, led by Health Data Research UK in partnership with the Office for National Statistics and funded by UK Research and Innovation (grant ref MC_PC_20058). We would like to express our sincere gratitude to the UK Biobank participants and administrative staff.

## Ethics Statement

The UK Biobank received ethical approval from the research ethics committee (reference ID: 11/ NW/0382). Written informed consent was obtained from all participants.

## Data availability statement

The datasets analyzed during the current study are available from the UK Biobank through an application process (www.ukbiobank.ac.uk/).

## Supplementary materials

Table S1. Characteristics of the participants by fish oil supplementation

Table S2. Demographic and clinical characteristics of the participants by fish oil supplementation after imputation

Table S3. New locus and known loci related to all-cause dementia, Alzheimer’s disease, and vascular dementia

Table S4. The risk of 81 SNPs in 43 loci and APOE ε4 genotype interacted with fish oil supplementation in the development of all-cause dementia in the whole participants.

Table S5. The risk of 81 SNPs in 43 loci and APOE ε4 genotype interacted with fish oil supplementation in the development of Alzheimer’s disease in the whole participants.

Table S6. The risk of 81 SNPs in 43 loci and APOE ε4 genotype interacted with fish oil supplementation in the development of vascular dementia in the whole participants.

Table S7. 43 Loci interacted with fish oil supplementation in the development of dementias compared to previous discoveries.

Table S8. Significance of interaction loci among GWAS in the whole dataset and subgroup dataset.

Table S9. Association between interaction loci and dementia outcomes in the whole dataset and fish oil supplementation subgroups.

Figure S1. Manhattan plots of GWAS of all-cause dementia by fish oil supplementation

Figure S2. Manhattan plots of GWAS of Alzheimer’s disease by fish oil supplementation

Figure S3. Manhattan plots of GWAS of vascular dementia by fish oil supplementation

Figure S4. Comparison of the associations between interaction loci and dementia outcomes stratified by fish oil supplementation.

Figure S5. Interaction analysis of candidate SNPs with fish oil supplementation in the development of dementia using Model 1-4.

Figure S6. Interaction analysis of candidate SNPs with fish oil supplementation from the touchscreen questionnaire and 24-hour recall questionnaire in the development of dementia by model 1.

Figure S7. Protein-coding genes enrich in tissues by adult and fetal status.

