## Supplementary Materials for "Gene-diet interaction analysis in UK Biobank identified genetic loci that modify the association between fish oil supplementation and the incidence of dementia"

### Supplementary Methods

#### *UK Biobank*

UK Biobank is a large-scale prospective study that recruited more than half a million people aged 40-69 who lived in the UK between 2006 and 2010.[1] Phenotypic and genotypic data were collected from questionnaires, interviews, physical measures, and biological samples at baseline. Follow-up data on health-related outcomes were accessed mainly through linkages to their records in available national datasets. Ethical approval was obtained from the North West Multi-centre Research Ethics Committee (reference 21/NW/0157). All participants provided written informed consent. This study was conducted using the UK Biobank resource (application number 48818).

#### *Sensitivity analysis*

##### *Exposure*

The touch-screen questionnaire completed at the assessment center provided information on fish oil supplementation intake as described in the Methods section. To evaluate the robustness of this questionnaire, we performed a sensitivity analysis on fish oil intake status based on the 24-hour dietary recall questionnaire. Participants were invited five times to repeatedly fill out this online questionnaire and were asked, "Did you have any vitamin or mineral supplements yesterday?". No more than half of the participants answered this questionnaire at least once. We divided all individuals into 24-hour recall groups and touch-screen groups in which people only completed the baseline questionnaire and performed time-to-event interaction analysis in two subgroups separately.

##### *Covariates*

For time-to-event GWAS, age, sex, and the top 10 genetic PCs were used as covariates. The same covariates, along with fish oil supplementation and the corresponding SNP, were included in model 1 for the gene × fish oil interaction analysis. Additionally, considering the potential effects caused by socioeconomic status, lifestyle, dietary patterns, and related medical history, several models adjusted by additional factors were performed as sensitivity analyses. Education (high/low), Townsend deprivation index (TDI), body mass index (BMI), smoking (never/previous/current), alcohol intake (non-drinker/low to moderate drinker/heavy drinker), and physical activity were included in model 2 on the basis of model 1. Model 3 further included 8 dietary patterns: oily fish intake (<1 times per week/1 times per week/≥2 times per week), fruit intake (<2 servings per day/2-3.9 servings per day/≥4 servings per day), vegetable intake (<2 servings per day/2-3.9 servings per day/≥4 servings per day), processed meat intake (<2 times per week/≥2 times per week), red meat intake (<2 times per week/≥2 times per week), vitamin supplementation (Yes/No), mineral supplementation (Yes/No), and Glucosamine supplementation (Yes/No). And 5 self-reported medical histories, hypertension, cardiovascular disease (CVD), high cholesterol, diabetes, and depression were included in model 4. The definitions of dietary patterns were described in the He Y., et al. study.[2] Self-reported medical histories were collected from verbal interviews at the time of recruitment. Specifically, CVD included angina, heart attack or myocardial infarction, stroke, subarachnoid haemorrhage, brain haemorrhage, and ischaemic stroke.

**Table S1.** Characteristics of the participants by fish oil supplementation.

| | Fish oil user (N = 113,267) | | Non fish oil user (N = 244,364) | | p-value |
|---|---|---|---|---|---|
| | Missing value, n | Values | Missing value, n | Values | |
| All-cause dementia, n (%) | | 2,613 (2.31) | | 4,345 (1.78) | <0.001 |
| Alzheimer's disease, n (%) | | 1,250 (1.10) | | 1,848 (0.76) | <0.001 |
| Vascular dementia, n (%) | | 542 (0.48) | | 1,015 (0.42) | 0.008 |
| Frontotemporal dementia, n (%) | | 57 (0.05) | | 162 (0.07) | 0.085 |
| Age, years | 0 | 58.94±7.29 | 0 | 55.82±8.14 | <0.001 |
| Female, n (%) | 0 | 63,514 (56.07) | 0 | 129,047 (52.81) | <0.001 |
| Body mass index, kg/m$^2$ | 312 | 27.15±4.51 | 843 | 27.49±4.87 | <0.001 |
| ***Socioeconomic factors*** | | | | | |
| Education, n (%) | 1,023 | | 1,970 | | <0.001 |
| High | | 51,008 (45.44) | | 119,909 (49.47) | |
| Low | | 61,236 (54.56) | | 122,485 (50.53) | |
| Townsend deprivation index | 97 | -2.48 [-3.81, -0.24] | 346 | -2.24 [-3.70, 0.32] | <0.001 |
| ***Lifestyle factors*** | | | | | |
| Smoking, n (%) | 415 | | 812 | | |
| Never | | 60,396 (53.52) | | 133,167 (54.68) | <0.001 |
| Previous | | 43,634 (38.66) | | 82,629 (33.93) | <0.001 |
| Current | | 8,822 (7.82) | | 27,756 (11.40) | <0.001 |
| Alcohol intake, n (%) | 74 | | 164 | | <0.001 |
| Non-drinker | | 18,537 (16.38) | | 42,392 (17.36) | |
| Low to moderate drinker | | 69,729 (61.60) | | 149,769 (61.33) | |
| Heavy drinker | | 24,927 (22.02) | | 52,039 (21.31) | |
| Physical activity, MET-min/week | 24,872 | 2,013.00 [956.00, 3,879.00] | 53,395 | 1,706.00 [773.00, 3,432.00] | <0.001 |
| ***Dietary patterns*** | | | | | |
| Oily fish intake, n (%) | 374 | | 1,205 | | <0.001 |
| <1 times/week | | 40,594 (35.96) | | 116,441 (47.89) | |
| 1 times/week | | 47,079 (41.70) | | 88,617 (36.44) | |
| ≥2 times/week | | 25,220 (22.34) | | 38,101 (15.67) | |
| Fruits, n (%) | 1,233 | | 2,744 | | <0.001 |
| <2 servings/day | | 31,481 (28.10) | | 93,942 (38.88) | |
| 2-3.9 servings/day | | 57,811 (51.60) | | 112,069 (46.38) | |
| ≥4 servings/day | | 22,742 (20.30) | | 35,609 (14.74) | |
| Vegetables, n (%) | 1,671 | | 4,204 | | <0.001 |
| <2 servings/day | | 73,797 (66.13) | | 170,422 (70.96) | |
| 2-3.9 servings/day | | 34,469 (30.89) | | 63,356 (26.38) | |
| ≥4 servings/day | | 3,330 (2.98) | | 6,382 (2.66) | |
| Processed meat, n (%) | 117 | | 373 | | |
| <2 times/week | | 80,457 (71.11) | | 163,525 (67.02) | <0.001 |
| ≥2 times/week | | 32,693 (28.89) | | 80,466 (32.98) | <0.001 |
| Red meats, n (%) | 693 | | 1,758 | | |

| | | | | | |
|---|---|---|---|---|---|
| <2 times/week | | 75,033 (66.65) | | 159,875 (65.90) | <0.001 |
| ≥2 times/week | | 37,541 (33.35) | | 82,731 (34.10) | <0.001 |
| Vitamins supplementation, n (%) | 609 | 63,050 (55.97) | 492 | 48,461 (19.87) | <0.001 |
| Minerals supplementation, n (%) | 0 | 23,432 (20.69) | 0 | 19,609 (8.02) | <0.001 |
| Glucosamine supplementation, n (%) | 0 | 44,179 (39.00) | 0 | 25,895 (10.60) | <0.001 |
| ***Self-reported medical histories*** | | | | | |
| Hypertension, n (%) | 0 | 31,669 (27.96) | 0 | 63,439 (25.96) | <0.001 |
| Cardiovascular disease, n (%) | 0 | 6,357 (5.61) | 0 | 13,630 (5.58) | 0.680 |
| High cholesterol, n (%) | 0 | 15,564 (13.74) | 0 | 27,981 (11.45) | <0.001 |
| Diabetes, n (%) | 0 | 4,905 (4.33) | 0 | 11,818 (4.84) | <0.001 |
| Depression, n (%) | 0 | 6,164 (5.44) | 0 | 14,524 (5.94) | <0.001 |

Normally distributed continuous variables were presented as mean ± standard deviation, non-normally distributed continuous variables were presented as median [interquartile range], and categorical variables were presented as frequency (percentage). And the difference between fish oil intake status groups was estimated by the Wilcoxon rank sum test or Chi-squared Test.

**Table S2.** Demographic and clinical characteristics of the participants by fish oil supplementation after imputation

| | **Fish oil user (N = 113,267)** | **Non-fish oil user (N = 244,364)** | **P-value** |
|---|---|---|---|
| Age, years | 58.94±7.29 | 55.82±8.14 | <0.001 |
| Female, n (%) | 63,514 (56.07) | 129,047 (52.81) | <0.001 |
| Education, n (%) | | | <0.001 |
| High | 51,443 (45.42) | 120,710 (49.40) | |
| Low | 61,824 (54.58) | 123,654 (50.60) | |
| Townsend deprivation index | -2.48 [-3.81, -0.24] | -2.24 [-3.70, 0.32] | <0.001 |
| Body mass index, kg/m$^2$ | 27.15±4.52 | 27.49±4.87 | <0.001 |
| Smoking, n (%) | | | |
| Never | 60,608 (53.51) | 133,589 (54.67) | <0.001 |
| Previous | 43,798 (38.67) | 82,937 (33.94) | <0.001 |
| Current | 8,861 (7.82) | 27,838 (11.39) | <0.001 |
| Alcohol intake, n (%) | | | <0.001 |
| Non-drinker | 18,550 (16.38) | 42,424 (17.36) | |
| Low to moderate drinker | 69,776 (61.60) | 149,861 (61.33) | |
| Heavy drinker | 24,941 (22.02) | 52,079 (21.31) | |
| Physical activity, MET-min/week | 1,986.00 [933.00, 3,828.00] | 1,710.00 [777.00, 3,450.00] | <0.001 |
| Oily fish intake, n (%) | | | <0.001 |
| <1 times/week | 40,768 (35.99) | 117,057 (47.90) | |
| 1 times/week | 47,213 (41.68) | 89,037 (36.44) | |
| ≥2 times/week | 25,286 (22.32) | 38,270 (15.66) | |
| Fruits, n (%) | | | <0.001 |
| <2 servings/day | 31,871 (28.14) | 95,029 (38.89) | |
| 2-3.9 servings/day | 58,413 (51.57) | 113,320 (46.37) | |
| ≥4 servings/day | 22,983 (20.29) | 36,015 (14.74) | |
| Vegetables, n (%) | | | <0.001 |
| <2 servings/day | 74,999 (66.21) | 173,494 (71.00) | |
| 2-3.9 servings/day | 34,899 (30.81) | 64,374 (26.34) | |
| ≥4 servings/day | 3,369 (2.97) | 6,496 (2.66) | |
| Processed meat, n (%) | | | |
| <2 times/week | 80,535 (71.10) | 163,755 (67.01) | <0.001 |
| ≥2 times/week | 32,732 (28.90) | 80,609 (32.99) | <0.001 |
| Red meats, n (%) | | | |
| <2 times/week | 75,492 (66.65) | 161,011 (65.89) | <0.001 |
| ≥2 times/week | 37,775 (33.35) | 83,353 (34.11) | <0.001 |
| Vitamins supplementation, n (%) | 63,290 (55.88) | 48,630 (19.90) | <0.001 |
| Minerals supplementation, n (%) | 23,432 (20.69) | 19,609 (8.02) | <0.001 |
| Glucosamine supplementation, n (%) | 44,179 (39.00) | 25,895 (10.60) | <0.001 |

Normally distributed continuous variables were presented as mean ± standard deviation, non-normally distributed continuous variables were presented as median [interquartile range], and categorical variables were presented as frequency (percentage). And the differences between fish oil intake status groups were estimated by Wilcoxon rank sum test or Chi-squared Test.

**Table S3.** The association between fish oil supplementation and all-cause dementia, Alzheimer's disease, and vascular dementia.

| | HR (95%CI) | P-value |
|---|---|---|
| ***All-cause dementia*** | | |
| Model 1 | 0.92 (0.87,0.96) | <0.001 |
| Model 2 | 0.95 (0.90,1.00) | 0.034 |
| Model 3 | 0.93 (0.88,0.98) | 0.009 |
| Model 4 | 0.94 (0.89,0.99) | 0.029 |
| ***Alzheimer's disease*** | | |
| Model 1 | 1.01 (0.94,1.08) | 0.863 |
| Model 2 | 1.03 (0.95,1.10) | 0.491 |
| Model 3 | 1.00 (0.92,1.08) | 0.916 |
| Model 4 | 1.00 (0.93,1.09) | 0.928 |
| ***Vascular dementia*** | | |
| Model 1 | 0.80 (0.72,0.89) | <0.001 |
| Model 2 | 0.85 (0.77,0.95) | 0.003 |
| Model 3 | 0.89 (0.80,1.00) | 0.058 |
| Model 4 | 0.92 (0.82,1.03) | 0.141 |

The associations between fish oil supplementation and dementia onsets were estimated using Cox regression models adjusted by age, sex, and 10 top genetic PCs.

Abbreviations: HR, hazard ratio; CI, confidence interval.

**Table S4.** New locus and known loci related to all-cause dementia, Alzheimer's disease, and vascular dementia.

| Outcome | No. | Chr | Start | End | Gene[a] | Known[b] (Yes/No) |
|---|---|---|---|---|---|---|
| All-cause dementia | 1 | 2 | 127597930 | 128144615 | *BIN1* | Yes |
| | 2 | 6 | 40654030 | 41379252 | *UNC5CL/TREML2/TREM2* | Yes |
| | 3 | 7 | 99527422 | 100234089 | *SPDYE3/ZCWPW1/NYAP1* | Yes |
| | 4 | 8 | 27206253 | 27718503 | *PTK2B/CLU* | Yes |
| | 5 | 11 | 59690599 | 60273087 | *MS4A4A/MS4A* | Yes |
| | 6 | 19 | 44893942 | 45961598 | *APOE* | Yes |
| Alzheimer's disease | 1 | 2 | 127641427 | 128141427 | *BIN1* | Yes |
| | 2 | 6 | 40654030 | 41379252 | *UNC5CL/TREML2/TREM2* | Yes |
| | 3 | 7 | 99527422 | 100234089 | *SPDYE3/ZCWPW1/NYAP1* | Yes |
| | 4 | 15 | 63319902 | 63821820 | *APH1B* | Yes |
| | 5 | 19 | 44882679 | 45925180 | *APOE* | Yes |
| Vascular dementia | 1 | 1 | 10672576 | 11174273 | *CASZ1* | No |
| | 2 | 19 | 45074138 | 45677125 | *APOE* | Yes |

[a]Gene symbols from the meta-analysis of GWAS on Alzheimer's disease and related dementias by Bellenguez, C. *et al.*[3] were used if the locus overlapped with their reports. The gene symbol for the unreported locus was identified as the nearest gene to the top SNPs within the locus by gene annotation from the Ensembl database.

[b]Known loci were identified as overlapped with the loci reported by Bellenguez, C. *et al.*

**Table S5.** Association between fish oil supplementation and the onset of all-cause dementia in each genotype of SNPs in 43 loci and *APOE* ε4 from the whole participants.

| CHR | No. Loci | Start | End | SNP | POS | REF | ALT | HR of fish oil supplementation in participants carrying REF allele[a] | | | | | | P-interaction |
|---|---|---|---|---|---|---|---|---|---|---|---|---|---|---|
| | | | | | | | | 0 | | 1 | | 2 | | |
| | | | | | | | | HR (95%CI) | P-value | HR (95%CI) | P-value | HR (95%CI) | P-value | |
| 19 | - | - | - | *APOE* ε4 | - | - | - | 0.88 (0.81,0.94) | <0.001 | 0.93 (0.87,1.01) | 0.082 | 0.97 (0.84,1.13) | 0.701 | 0.029 |
| 1 | 1 | 8027198 | 8527198 | rs116501531 | 8277198 | A | G | 0.91 (0.86,0.95) | <0.001 | 1.36 (1.03,1.81) | 0.031 | - | - | 0.007 |
| 1 | 2 | 14816263 | 15316263 | rs75837905 | 15066263 | A | G | 0.89 (0.85,0.94) | <0.001 | 1.44 (1.16,1.80) | 0.001 | - | - | <0.001 |
| 1 | 3 | 29232654 | 29732654 | rs116264291 | 29482654 | A | G | 0.90 (0.86,0.95) | <0.001 | 1.42 (1.11,1.83) | 0.006 | - | - | <0.001 |
| 1 | 4 | 101852308 | 102352308 | rs61804494 | 102102308 | G | A | 0.97 (0.91,1.03) | 0.304 | 0.82 (0.74,0.90) | <0.001 | 0.62 (0.45,0.86) | 0.004 | <0.001 |
| 1 | 5 | 106483537 | 106983795 | rs12043527 | 106733537 | G | A | 0.87 (0.83,0.92) | <0.001 | 1.10 (0.98,1.23) | 0.108 | 1.20 (0.79,1.83) | 0.395 | <0.001 |
| 1 | 5 | 106483537 | 106983795 | rs11184799 | 106733795 | G | A | 0.88 (0.83,0.93) | <0.001 | 1.05 (0.95,1.17) | 0.316 | 1.12 (0.80,1.57) | 0.526 | <0.001 |
| 1 | 6 | 110597781 | 111098352 | rs12024138 | 110847781 | C | T | 0.88 (0.84,0.93) | <0.001 | 1.29 (1.10,1.52) | 0.002 | 1.43 (0.52,3.92) | 0.485 | <0.001 |
| 1 | 6 | 110597781 | 111098352 | rs12024264 | 110848352 | C | A | 0.88 (0.84,0.93) | <0.001 | 1.29 (1.10,1.52) | 0.002 | 1.43 (0.52,3.92) | 0.485 | <0.001 |
| 1 | 7 | 222375645 | 222876659 | rs17163136 | 222625645 | T | C | 0.99 (0.92,1.07) | 0.829 | 0.90 (0.84,0.97) | 0.006 | 0.77 (0.68,0.88) | <0.001 | 0.002 |
| 1 | 7 | 222375645 | 222876659 | rs17163137 | 222626659 | T | C | 0.99 (0.92,1.07) | 0.831 | 0.90 (0.84,0.97) | 0.005 | 0.78 (0.69,0.89) | <0.001 | 0.003 |
| 2 | 8 | 100571438 | 101071438 | rs113777826 | 100821438 | C | T | 0.91 (0.86,0.95) | <0.001 | 1.28 (0.97,1.70) | 0.08 | - | - | 0.017 |
| 2 | 9 | 170757664 | 171257664 | rs4435418 | 171007664 | C | T | 0.81 (0.73,0.91) | <0.001 | 0.89 (0.83,0.95) | <0.001 | 1.05 (0.96,1.15) | 0.284 | <0.001 |
| 4 | 10 | 16312945 | 16812945 | rs148811174 | 16562945 | C | T | 0.98 (0.92,1.04) | 0.421 | 0.79 (0.73,0.87) | <0.001 | 0.94 (0.73,1.21) | 0.651 | 0.003 |
| 4 | 11 | 68145282 | 68645282 | rs149325653 | 68395282 | A | G | 0.93 (0.88,0.97) | 0.003 | 0.66 (0.48,0.91) | 0.011 | - | - | 0.037 |
| 4 | 12 | 114132769 | 114635835 | rs13123494 | 114382769 | C | A | 0.85 (0.79,0.92) | <0.001 | 0.93 (0.86,0.99) | 0.036 | 1.03 (0.91,1.17) | 0.623 | 0.009 |
| 4 | 12 | 114132769 | 114635835 | rs1525000 | 114384869 | C | T | 0.85 (0.78,0.92) | <0.001 | 0.93 (0.87,1.00) | 0.048 | 1.02 (0.90,1.15) | 0.794 | 0.011 |
| 4 | 12 | 114132769 | 114635835 | rs62314976 | 114385676 | T | C | 0.85 (0.78,0.92) | <0.001 | 0.93 (0.87,1.00) | 0.054 | 1.02 (0.90,1.16) | 0.72 | 0.008 |

| | | | | | | | | | | | | | | |
|---|---|---|---|---|---|---|---|---|---|---|---|---|---|---|
| 4 | 12 | 114132769 | 114635835 | rs13106836 | 114385835 | G | A | 0.85 (0.78,0.92) | <0.001 | 0.93 (0.87,1.00) | 0.054 | 1.02 (0.90,1.15) | 0.734 | 0.008 |
| 5 | 13 | 3009680 | 3517512 | rs7722735 | 3259680 | G | T | 0.96 (0.90,1.03) | 0.268 | 0.91 (0.85,0.98) | 0.016 | 0.73 (0.62,0.86) | <0.001 | 0.006 |
| 5 | 13 | 3009680 | 3517512 | rs59781823 | 3262724 | G | A | 0.96 (0.89,1.03) | 0.265 | 0.91 (0.85,0.98) | 0.018 | 0.72 (0.61,0.85) | <0.001 | 0.004 |
| 5 | 13 | 3009680 | 3517512 | rs60808439 | 3267512 | C | T | 0.95 (0.89,1.01) | 0.088 | 0.89 (0.82,0.97) | 0.008 | 0.73 (0.57,0.93) | 0.012 | 0.052 |
| 5 | 14 | 100147117 | 100647117 | rs2089903 | 100397117 | C | A | - | - | 1.39 (1.10,1.74) | 0.005 | 0.90 (0.85,0.94) | <0.001 | <0.001 |
| 6 | 15 | 767009 | 1267009 | rs845890 | 1017009 | C | A | 0.89 (0.84,0.95) | <0.001 | 0.95 (0.87,1.03) | 0.21 | 0.98 (0.77,1.24) | 0.844 | 0.238 |
| 6 | 16 | 115069829 | 115569829 | rs9505681 | 115319829 | C | T | 0.90 (0.85,0.94) | <0.001 | 1.61 (1.21,2.15) | 0.001 | - | - | <0.001 |
| 6 | 17 | 166801161 | 167302048 | rs6941412 | 167051161 | C | T | 1.01 (0.94,1.07) | 0.881 | 0.85 (0.78,0.92) | <0.001 | 0.70 (0.57,0.84) | <0.001 | <0.001 |
| 6 | 17 | 166801161 | 167302048 | rs12174679 | 167051368 | G | A | 1.00 (0.94,1.07) | 0.885 | 0.84 (0.78,0.91) | <0.001 | 0.71 (0.58,0.87) | <0.001 | <0.001 |
| 6 | 17 | 166801161 | 167302048 | rs963302 | 167052048 | C | G | 1.01 (0.94,1.08) | 0.859 | 0.86 (0.79,0.93) | <0.001 | 0.68 (0.57,0.82) | <0.001 | <0.001 |
| 7 | 18 | 20096169 | 20596169 | rs3114430 | 20346169 | C | A | 1.13 (0.98,1.30) | 0.088 | 0.95 (0.88,1.03) | 0.192 | 0.81 (0.75,0.88) | <0.001 | <0.001 |
| 7 | 19 | 143574310 | 144074310 | rs12670543 | 143824310 | C | A | 0.86 (0.82,0.91) | <0.001 | 1.17 (1.04,1.31) | 0.007 | 1.23 (0.74,2.04) | 0.417 | <0.001 |
| 8 | 20 | 3675912 | 4175912 | rs2042528 | 3925912 | A | T | 0.99 (0.90,1.09) | 0.833 | 0.93 (0.87,1.00) | 0.058 | 0.80 (0.72,0.89) | <0.001 | 0.005 |
| 8 | 21 | 82617116 | 83117116 | rs117216345 | 82867116 | T | A | 0.91 (0.86,0.96) | <0.001 | 0.99 (0.87,1.13) | 0.865 | 1.00 (0.55,1.85) | 0.989 | 0.242 |
| 8 | 22 | 111221071 | 111721071 | rs116871946 | 111471071 | T | C | 0.93 (0.89,0.98) | 0.009 | 0.76 (0.64,0.90) | 0.001 | 0.58 (0.25,1.36) | 0.211 | 0.01 |
| 10 | 23 | 4149849 | 4649849 | rs116876958 | 4399849 | C | T | 0.91 (0.86,0.96) | <0.001 | 1.03 (0.84,1.26) | 0.785 | - | - | 0.165 |
| 10 | 24 | 129615006 | 130115006 | rs4750683 | 129865006 | G | C | 1.05 (0.87,1.26) | 0.61 | 1.02 (0.94,1.11) | 0.593 | 0.84 (0.79,0.90) | <0.001 | <0.001 |
| 11 | 25 | 85731587 | 86233703 | rs75559794 | 85981587 | G | A | 0.89 (0.85,0.94) | <0.001 | 1.06 (0.93,1.21) | 0.363 | 1.26 (0.67,2.37) | 0.471 | 0.01 |
| 11 | 25 | 85731587 | 86233703 | rs78851816 | 85983703 | G | A | 0.89 (0.85,0.94) | <0.001 | 1.06 (0.93,1.21) | 0.394 | 1.28 (0.68,2.40) | 0.45 | 0.01 |
| 11 | 26 | 96280700 | 96825966 | rs754413 | 96530700 | A | G | 1.54 (1.03,2.29) | 0.035 | 1.04 (0.93,1.15) | 0.512 | 0.88 (0.83,0.93) | <0.001 | <0.001 |
| 11 | 26 | 96280700 | 96825966 | rs3018644 | 96575966 | G | A | 1.45 (0.96,2.18) | 0.077 | 1.06 (0.95,1.17) | 0.287 | 0.87 (0.82,0.92) | <0.001 | <0.001 |
| 12 | 27 | 47212480 | 47712480 | rs74523587 | 47462480 | A | C | 0.90 (0.85,0.94) | <0.001 | 1.40 (1.12,1.76) | 0.003 | - | - | <0.001 |
| 12 | 28 | 55968568 | 56468568 | rs73119275 | 56218568 | C | T | 0.90 (0.85,0.94) | <0.001 | 1.56 (1.22,2.00) | <0.001 | - | - | <0.001 |

| | | | | | | | | | | | | | | |
|---|---|---|---|---|---|---|---|---|---|---|---|---|---|---|
| 12 | 29 | 69525118 | 70042102 | rs2870901 | 69775118 | C | T | 0.80 (0.68,0.94) | 0.006 | 0.90 (0.83,0.97) | 0.004 | 0.96 (0.89,1.03) | 0.21 | 0.024 |
| 12 | 29 | 69525118 | 70042102 | rs1585705 | 69792102 | A | C | 0.77 (0.65,0.90) | <0.001 | 0.91 (0.84,0.98) | 0.011 | 0.96 (0.89,1.03) | 0.265 | 0.012 |
| 12 | 30 | 104063861 | 104572214 | rs11111842 | 104313861 | A | G | 0.91 (0.86,0.96) | <0.001 | 0.96 (0.83,1.10) | 0.546 | 1.13 (0.44,2.88) | 0.803 | 0.494 |
| 12 | 30 | 104063861 | 104572214 | rs10861145 | 104314531 | G | C | 0.91 (0.86,0.96) | <0.001 | 0.96 (0.83,1.10) | 0.545 | 1.13 (0.44,2.88) | 0.803 | 0.495 |
| 12 | 30 | 104063861 | 104572214 | rs11833702 | 104314560 | A | G | 0.91 (0.86,0.96) | <0.001 | 0.96 (0.84,1.11) | 0.58 | 1.13 (0.44,2.88) | 0.805 | 0.463 |
| 12 | 30 | 104063861 | 104572214 | rs11111843 | 104314956 | T | C | 0.91 (0.86,0.96) | <0.001 | 0.96 (0.83,1.10) | 0.545 | 1.13 (0.44,2.88) | 0.803 | 0.498 |
| 12 | 30 | 104063861 | 104572214 | rs11111844 | 104316005 | T | C | 0.91 (0.86,0.96) | <0.001 | 0.96 (0.84,1.10) | 0.566 | 1.13 (0.44,2.88) | 0.804 | 0.477 |
| 12 | 30 | 104063861 | 104572214 | rs76025677 | 104317050 | A | G | 0.91 (0.86,0.96) | <0.001 | 0.96 (0.84,1.11) | 0.6 | 1.13 (0.44,2.88) | 0.804 | 0.45 |
| 12 | 30 | 104063861 | 104572214 | rs11111846 | 104318346 | T | C | 0.91 (0.86,0.96) | <0.001 | 0.96 (0.84,1.10) | 0.566 | 1.13 (0.44,2.88) | 0.804 | 0.47 |
| 12 | 30 | 104063861 | 104572214 | rs11111849 | 104319873 | A | G | 0.91 (0.86,0.96) | <0.001 | 0.96 (0.84,1.10) | 0.566 | 1.13 (0.44,2.88) | 0.804 | 0.475 |
| 12 | 30 | 104063861 | 104572214 | rs11111850 | 104321073 | A | T | 0.91 (0.86,0.96) | <0.001 | 0.96 (0.84,1.10) | 0.566 | 1.13 (0.44,2.88) | 0.804 | 0.474 |
| 12 | 30 | 104063861 | 104572214 | rs3794246 | 104321902 | T | C | 0.91 (0.86,0.96) | <0.001 | 0.96 (0.83,1.10) | 0.541 | 1.13 (0.44,2.89) | 0.8 | 0.495 |
| 12 | 30 | 104063861 | 104572214 | rs17034916 | 104322214 | C | T | 0.91 (0.86,0.96) | <0.001 | 0.96 (0.84,1.10) | 0.566 | 1.13 (0.44,2.88) | 0.804 | 0.474 |
| 13 | 31 | 26140990 | 26728255 | rs117674980 | 26390990 | A | G | 0.92 (0.88,0.97) | 0.001 | 0.83 (0.68,1.01) | 0.056 | 0.31 (0.06,1.62) | 0.165 | 0.186 |
| 13 | 31 | 26140990 | 26728255 | rs77999054 | 26394850 | G | A | 0.92 (0.88,0.97) | 0.001 | 0.84 (0.69,1.02) | 0.072 | 0.31 (0.06,1.62) | 0.165 | 0.222 |
| 13 | 31 | 26140990 | 26728255 | rs3783124 | 26458231 | C | T | 0.92 (0.88,0.97) | 0.001 | 0.85 (0.70,1.04) | 0.108 | 0.06 (0.00,2.91) | 0.153 | 0.27 |
| 13 | 31 | 26140990 | 26728255 | rs74335017 | 26478255 | C | T | 0.92 (0.88,0.97) | 0.001 | 0.87 (0.72,1.06) | 0.172 | 0.03 (0.00,1.41) | 0.075 | 0.359 |
| 13 | 32 | 67331127 | 67831127 | rs9571707 | 67581127 | A | G | 0.85 (0.79,0.91) | <0.001 | 0.96 (0.89,1.04) | 0.313 | 1.11 (0.93,1.31) | 0.247 | <0.001 |
| 13 | 33 | 93315090 | 93875433 | rs1932193 | 93565090 | A | C | 0.85 (0.78,0.93) | <0.001 | 0.95 (0.89,1.02) | 0.179 | 0.95 (0.85,1.06) | 0.364 | 0.081 |
| 13 | 33 | 93315090 | 93875433 | rs306675 | 93606083 | A | C | 0.83 (0.74,0.93) | 0.002 | 0.94 (0.88,1.01) | 0.092 | 0.93 (0.86,1.02) | 0.125 | 0.161 |
| 13 | 33 | 93315090 | 93875433 | rs306677 | 93613362 | C | G | 0.83 (0.74,0.94) | 0.003 | 0.94 (0.88,1.01) | 0.087 | 0.94 (0.86,1.02) | 0.134 | 0.169 |
| 13 | 33 | 93315090 | 93875433 | rs306679 | 93625433 | G | T | 0.84 (0.74,0.95) | 0.006 | 0.95 (0.88,1.02) | 0.157 | 0.93 (0.85,1.01) | 0.08 | 0.302 |
| 13 | 34 | 100252104 | 100752104 | rs72653992 | 100502104 | A | G | 0.90 (0.86,0.95) | <0.001 | 1.10 (0.92,1.32) | 0.298 | 0.73 (0.22,2.44) | 0.61 | 0.039 |

| | | | | | | | | | | | | | | |
|---|---|---|---|---|---|---|---|---|---|---|---|---|---|---|
| 14 | 35 | 57055847 | 57555847 | rs12896185 | 57305847 | A | G | 0.85 (0.80,0.91) | <0.001 | 0.99 (0.91,1.08) | 0.829 | 1.23 (1.00,1.51) | 0.052 | <0.001 |
| 14 | 36 | 80321393 | 80823172 | rs1181351 | 80571393 | G | T | 0.86 (0.71,1.03) | 0.094 | 0.95 (0.88,1.02) | 0.169 | 0.89 (0.83,0.96) | 0.001 | 0.657 |
| 14 | 36 | 80321393 | 80823172 | rs28444185 | 80573172 | A | T | 0.89 (0.83,0.95) | <0.001 | 0.95 (0.88,1.03) | 0.204 | 0.86 (0.72,1.03) | 0.103 | 0.575 |
| 15 | 37 | 90077226 | 90587639 | rs28698386 | 90327226 | T | C | 1.01 (0.91,1.11) | 0.884 | 0.91 (0.85,0.97) | 0.007 | 0.86 (0.78,0.95) | 0.002 | 0.02 |
| 15 | 37 | 90077226 | 90587639 | rs6496605 | 90337639 | C | T | 1.00 (0.91,1.11) | 0.939 | 0.90 (0.84,0.97) | 0.004 | 0.85 (0.77,0.94) | 0.001 | 0.016 |
| 16 | 38 | 13669429 | 14169429 | rs75422462 | 13919429 | G | A | 0.91 (0.86,0.95) | <0.001 | 1.17 (0.91,1.51) | 0.226 | - | - | 0.029 |
| 18 | 39 | 52608233 | 53108233 | rs34352315 | 52858233 | G | A | 0.93 (0.89,0.98) | 0.004 | 0.53 (0.38,0.73) | <0.001 | - | - | <0.001 |
| 21 | 40 | 17158456 | 17658456 | rs2823575 | 17408456 | G | A | 0.89 (0.84,0.94) | <0.001 | 1.05 (0.94,1.17) | 0.408 | 0.63 (0.39,1.03) | 0.064 | 0.099 |
| 21 | 41 | 40203038 | 40703038 | rs74482819 | 40453038 | C | T | 0.90 (0.86,0.95) | <0.001 | 1.33 (1.04,1.69) | 0.021 | - | - | 0.004 |
| 22 | 42 | 18315772 | 18842083 | rs71328255 | 18565772 | A | G | 0.90 (0.85,0.94) | <0.001 | 1.57 (1.21,2.05) | <0.001 | - | - | <0.001 |
| 22 | 42 | 18315772 | 18842083 | rs71328256 | 18569926 | T | C | 0.90 (0.85,0.94) | <0.001 | 1.57 (1.21,2.05) | <0.001 | - | - | <0.001 |
| 22 | 42 | 18315772 | 18842083 | rs34680388 | 18576176 | T | A | 0.90 (0.85,0.94) | <0.001 | 1.57 (1.21,2.04) | <0.001 | - | - | <0.001 |
| 22 | 42 | 18315772 | 18842083 | rs13058445 | 18580087 | A | G | 0.90 (0.85,0.94) | <0.001 | 1.57 (1.20,2.04) | <0.001 | - | - | <0.001 |
| 22 | 42 | 18315772 | 18842083 | rs34658760 | 18584324 | A | C | 0.90 (0.85,0.94) | <0.001 | 1.58 (1.21,2.06) | <0.001 | - | - | <0.001 |
| 22 | 42 | 18315772 | 18842083 | rs34667409 | 18584379 | A | G | 0.90 (0.85,0.94) | <0.001 | 1.57 (1.21,2.04) | <0.001 | - | - | <0.001 |
| 22 | 42 | 18315772 | 18842083 | rs35137695 | 18587898 | C | T | 0.90 (0.85,0.94) | <0.001 | 1.59 (1.22,2.07) | <0.001 | - | - | <0.001 |
| 22 | 42 | 18315772 | 18842083 | rs13058179 | 18592083 | G | A | 0.90 (0.85,0.94) | <0.001 | 1.54 (1.18,2.00) | 0.001 | - | - | <0.001 |
| 22 | 43 | 29569106 | 30069106 | rs144548824 | 29819106 | G | A | 0.94 (0.89,0.99) | 0.011 | 0.59 (0.47,0.75) | <0.001 | - | - | <0.001 |

[a]The risk of fish oil supplementation for the development of all-cause dementia was evaluated using Cox regression models adjusted by age, sex, and 10 top genetic principal components within each SNP genotype. Genotype groups with five or fewer incident cases were not considered in the subgroup analysis.

Abbreviations: CHR, chromosome; SNP, single nucleotide polymorphism; POS, SNP position with GRCh37 assembly; REF, reference allele; ALT, alternative allele; HR, hazard ratio; CI, confidence interval; P-interaction, P value of the interaction terms.

**Table S6.** Association between fish oil supplementation and the onset of Alzheimer's disease in each genotype of SNPs in 43 loci and *APOE* ε4 from the whole participants.

| CHR | No. Loci | Start | End | SNP | POS | REF | ALT | HR of fish oil supplementation in participants carrying REF allele[a] | | | | | | P-interaction |
|---|---|---|---|---|---|---|---|---|---|---|---|---|---|---|
| | | | | | | | | 0 | | 1 | | 2 | | |
| | | | | | | | | HR (95%CI) | P-value | HR (95%CI) | P-value | HR (95%CI) | P-value | |
| 19 | - | - | - | *APOE* ε4 | - | - | - | 1.01 (0.90,1.14) | 0.821 | 0.99 (0.89,1.10) | 0.911 | 1.03 (0.85,1.25) | 0.752 | 0.912 |
| 1 | 1 | 8027198 | 8527198 | rs116501531 | 8277198 | A | G | 0.98 (0.91,1.05) | 0.57 | 2.18 (1.48,3.20) | <0.001 | - | - | <0.001 |
| 1 | 2 | 14816263 | 15316263 | rs75837905 | 15066263 | A | G | 0.97 (0.90,1.04) | 0.42 | 1.95 (1.42,2.66) | <0.001 | - | - | <0.001 |
| 1 | 3 | 29232654 | 29732654 | rs116264291 | 29482654 | A | G | 0.99 (0.92,1.07) | 0.815 | 1.58 (1.06,2.35) | 0.026 | - | - | 0.012 |
| 1 | 4 | 101852308 | 102352308 | rs61804494 | 102102308 | G | A | 1.08 (0.99,1.18) | 0.069 | 0.85 (0.74,0.98) | 0.029 | 0.50 (0.28,0.89) | 0.018 | <0.001 |
| 1 | 5 | 106483537 | 106983795 | rs12043527 | 106733537 | G | A | 0.97 (0.90,1.05) | 0.479 | 1.17 (0.99,1.40) | 0.073 | 1.51 (0.82,2.78) | 0.182 | 0.012 |
| 1 | 5 | 106483537 | 106983795 | rs11184799 | 106733795 | G | A | 0.99 (0.91,1.08) | 0.811 | 1.10 (0.94,1.29) | 0.214 | 1.36 (0.85,2.17) | 0.206 | 0.076 |
| 1 | 6 | 110597781 | 111098352 | rs12024138 | 110847781 | C | T | 0.96 (0.89,1.03) | 0.271 | 1.63 (1.28,2.06) | <0.001 | 1.44 (0.31,6.57) | 0.639 | <0.001 |
| 1 | 6 | 110597781 | 111098352 | rs12024264 | 110848352 | C | A | 0.96 (0.89,1.03) | 0.271 | 1.63 (1.28,2.06) | <0.001 | 1.44 (0.31,6.57) | 0.639 | <0.001 |
| 1 | 7 | 222375645 | 222876659 | rs17163136 | 222625645 | T | C | 1.08 (0.96,1.21) | 0.212 | 1.01 (0.91,1.12) | 0.873 | 0.83 (0.69,1.01) | 0.068 | 0.056 |
| 1 | 7 | 222375645 | 222876659 | rs17163137 | 222626659 | T | C | 1.08 (0.96,1.21) | 0.219 | 1.01 (0.91,1.12) | 0.883 | 0.84 (0.69,1.02) | 0.079 | 0.065 |
| 2 | 8 | 100571438 | 101071438 | rs113777826 | 100821438 | C | T | 1.01 (0.94,1.09) | 0.77 | 1.04 (0.67,1.63) | 0.86 | - | - | 0.851 |
| 2 | 9 | 170757664 | 171257664 | rs4435418 | 171007664 | C | T | 0.93 (0.79,1.10) | 0.392 | 0.96 (0.87,1.07) | 0.45 | 1.14 (1.00,1.30) | 0.057 | 0.038 |
| 4 | 10 | 16312945 | 16812945 | rs148811174 | 16562945 | C | T | 1.03 (0.94,1.13) | 0.485 | 0.93 (0.82,1.06) | 0.303 | 1.19 (0.82,1.72) | 0.364 | 0.675 |
| 4 | 11 | 68145282 | 68645282 | rs149325653 | 68395282 | A | G | 1.04 (0.97,1.12) | 0.25 | 0.40 (0.24,0.65) | <0.001 | - | - | <0.001 |
| 4 | 12 | 114132769 | 114635835 | rs13123494 | 114382769 | C | A | 1.01 (0.89,1.13) | 0.921 | 1.01 (0.91,1.12) | 0.862 | 1.02 (0.85,1.23) | 0.801 | 0.874 |
| 4 | 12 | 114132769 | 114635835 | rs1525000 | 114384869 | C | T | 1.00 (0.89,1.13) | 0.988 | 1.01 (0.91,1.13) | 0.806 | 1.02 (0.85,1.22) | 0.871 | 0.881 |
| 4 | 12 | 114132769 | 114635835 | rs62314976 | 114385676 | T | C | 0.99 (0.88,1.12) | 0.916 | 1.02 (0.92,1.14) | 0.691 | 1.01 (0.84,1.21) | 0.916 | 0.811 |

| | | | | | | | | | | | | | | |
|---|---|---|---|---|---|---|---|---|---|---|---|---|---|---|
| 4 | 12 | 114132769 | 114635835 | rs13106836 | 114385835 | G | A | 0.99 (0.88,1.12) | 0.924 | 1.02 (0.92,1.14) | 0.694 | 1.01 (0.84,1.21) | 0.915 | 0.813 |
| 5 | 13 | 3009680 | 3517512 | rs7722735 | 3259680 | G | T | 1.14 (1.03,1.27) | 0.013 | 0.96 (0.86,1.07) | 0.495 | 0.69 (0.55,0.88) | 0.002 | <0.001 |
| 5 | 13 | 3009680 | 3517512 | rs59781823 | 3262724 | G | A | 1.14 (1.03,1.27) | 0.014 | 0.96 (0.86,1.08) | 0.511 | 0.68 (0.54,0.87) | 0.002 | <0.001 |
| 5 | 13 | 3009680 | 3517512 | rs60808439 | 3267512 | C | T | 1.12 (1.02,1.23) | 0.023 | 0.91 (0.80,1.03) | 0.119 | 0.62 (0.43,0.89) | 0.01 | <0.001 |
| 5 | 14 | 100147117 | 100647117 | rs2089903 | 100397117 | C | A | - | - | 1.56 (1.11,2.19) | 0.011 | 0.99 (0.92,1.06) | 0.763 | 0.029 |
| 6 | 15 | 767009 | 1267009 | rs845890 | 1017009 | C | A | 0.99 (0.90,1.08) | 0.814 | 1.03 (0.90,1.17) | 0.67 | 1.10 (0.75,1.60) | 0.627 | 0.389 |
| 6 | 16 | 115069829 | 115569829 | rs9505681 | 115319829 | C | T | 0.98 (0.91,1.06) | 0.667 | 2.08 (1.31,3.29) | 0.002 | - | - | 0.001 |
| 6 | 17 | 166801161 | 167302048 | rs6941412 | 167051161 | C | T | 1.08 (0.98,1.19) | 0.118 | 0.96 (0.86,1.09) | 0.541 | 0.75 (0.56,0.99) | 0.043 | 0.006 |
| 6 | 17 | 166801161 | 167302048 | rs12174679 | 167051368 | G | A | 1.09 (0.99,1.20) | 0.087 | 0.94 (0.84,1.06) | 0.344 | 0.79 (0.59,1.06) | 0.122 | 0.007 |
| 6 | 17 | 166801161 | 167302048 | rs963302 | 167052048 | C | G | 1.08 (0.98,1.19) | 0.107 | 0.97 (0.86,1.09) | 0.575 | 0.73 (0.55,0.97) | 0.027 | 0.004 |
| 7 | 18 | 20096169 | 20596169 | rs3114430 | 20346169 | C | A | 1.20 (0.97,1.48) | 0.092 | 1.00 (0.90,1.12) | 0.983 | 0.95 (0.85,1.06) | 0.339 | 0.078 |
| 7 | 19 | 143574310 | 144074310 | rs12670543 | 143824310 | C | A | 0.96 (0.88,1.04) | 0.269 | 1.24 (1.05,1.48) | 0.013 | 1.29 (0.58,2.89) | 0.53 | 0.008 |
| 8 | 20 | 3675912 | 4175912 | rs2042528 | 3925912 | A | T | 1.23 (1.07,1.41) | 0.004 | 1.01 (0.91,1.12) | 0.866 | 0.80 (0.68,0.94) | 0.007 | <0.001 |
| 8 | 21 | 82617116 | 83117116 | rs117216345 | 82867116 | T | A | 1.03 (0.95,1.12) | 0.431 | 0.97 (0.80,1.18) | 0.753 | 0.64 (0.20,2.00) | 0.439 | 0.334 |
| 8 | 22 | 111221071 | 111721071 | rs116871946 | 111471071 | T | C | 1.05 (0.97,1.13) | 0.198 | 0.68 (0.53,0.87) | 0.002 | 0.10 (0.01,0.85) | 0.035 | <0.001 |
| 10 | 23 | 4149849 | 4649849 | rs116876958 | 4399849 | C | T | 1.01 (0.94,1.09) | 0.806 | 0.96 (0.70,1.33) | 0.819 | - | - | 0.991 |
| 10 | 24 | 129615006 | 130115006 | rs4750683 | 129865006 | G | C | 1.14 (0.87,1.49) | 0.334 | 1.16 (1.03,1.31) | 0.015 | 0.91 (0.82,1.00) | 0.056 | 0.005 |
| 11 | 25 | 85731587 | 86233703 | rs75559794 | 85981587 | G | A | 0.94 (0.87,1.02) | 0.118 | 1.48 (1.23,1.79) | <0.001 | 1.48 (0.66,3.35) | 0.342 | <0.001 |
| 11 | 25 | 85731587 | 86233703 | rs78851816 | 85983703 | G | A | 0.94 (0.87,1.01) | 0.11 | 1.49 (1.24,1.79) | <0.001 | 1.50 (0.67,3.39) | 0.328 | <0.001 |
| 11 | 26 | 96280700 | 96825966 | rs754413 | 96530700 | A | G | 1.46 (0.83,2.57) | 0.19 | 1.20 (1.03,1.41) | 0.022 | 0.95 (0.88,1.03) | 0.254 | 0.003 |
| 11 | 26 | 96280700 | 96825966 | rs3018644 | 96575966 | G | A | 1.17 (0.64,2.15) | 0.602 | 1.22 (1.04,1.42) | 0.013 | 0.95 (0.88,1.04) | 0.259 | 0.006 |
| 12 | 27 | 47212480 | 47712480 | rs74523587 | 47462480 | A | C | 0.99 (0.92,1.07) | 0.882 | 1.42 (1.02,1.97) | 0.037 | - | - | 0.051 |
| 12 | 28 | 55968568 | 56468568 | rs73119275 | 56218568 | C | T | 0.98 (0.91,1.06) | 0.671 | 1.86 (1.28,2.70) | 0.001 | - | - | <0.001 |

| | | | | | | | | | | | | | | |
|---|---|---|---|---|---|---|---|---|---|---|---|---|---|---|
| 12 | 29 | 69525118 | 70042102 | rs2870901 | 69775118 | C | T | 0.90 (0.71,1.13) | 0.364 | 1.04 (0.93,1.17) | 0.467 | 1.00 (0.90,1.11) | 0.998 | 0.634 |
| 12 | 29 | 69525118 | 70042102 | rs1585705 | 69792102 | A | C | 0.78 (0.62,0.99) | 0.038 | 1.04 (0.93,1.16) | 0.504 | 1.03 (0.93,1.14) | 0.582 | 0.116 |
| 12 | 30 | 104063861 | 104572214 | rs11111842 | 104313861 | A | G | 1.01 (0.93,1.09) | 0.837 | 1.02 (0.83,1.25) | 0.854 | 0.40 (0.04,3.56) | 0.409 | 0.887 |
| 12 | 30 | 104063861 | 104572214 | rs10861145 | 104314531 | G | C | 1.01 (0.93,1.09) | 0.836 | 1.02 (0.83,1.25) | 0.855 | 0.40 (0.04,3.56) | 0.409 | 0.886 |
| 12 | 30 | 104063861 | 104572214 | rs11833702 | 104314560 | A | G | 1.01 (0.93,1.09) | 0.826 | 1.02 (0.83,1.25) | 0.857 | 0.40 (0.04,3.56) | 0.409 | 0.882 |
| 12 | 30 | 104063861 | 104572214 | rs11111843 | 104314956 | T | C | 1.01 (0.93,1.09) | 0.825 | 1.02 (0.83,1.25) | 0.855 | 0.40 (0.04,3.56) | 0.409 | 0.881 |
| 12 | 30 | 104063861 | 104572214 | rs11111844 | 104316005 | T | C | 1.01 (0.93,1.09) | 0.825 | 1.02 (0.83,1.25) | 0.852 | 0.40 (0.04,3.56) | 0.409 | 0.884 |
| 12 | 30 | 104063861 | 104572214 | rs76025677 | 104317050 | A | G | 1.01 (0.93,1.09) | 0.835 | 1.02 (0.83,1.26) | 0.824 | 0.40 (0.04,3.56) | 0.409 | 0.915 |
| 12 | 30 | 104063861 | 104572214 | rs11111846 | 104318346 | T | C | 1.01 (0.93,1.09) | 0.855 | 1.02 (0.83,1.25) | 0.851 | 0.40 (0.04,3.56) | 0.409 | 0.896 |
| 12 | 30 | 104063861 | 104572214 | rs11111849 | 104319873 | A | G | 1.01 (0.93,1.09) | 0.836 | 1.02 (0.83,1.25) | 0.852 | 0.40 (0.04,3.56) | 0.409 | 0.889 |
| 12 | 30 | 104063861 | 104572214 | rs11111850 | 104321073 | A | T | 1.01 (0.93,1.09) | 0.836 | 1.02 (0.83,1.25) | 0.852 | 0.40 (0.04,3.56) | 0.409 | 0.889 |
| 12 | 30 | 104063861 | 104572214 | rs3794246 | 104321902 | T | C | 1.01 (0.93,1.09) | 0.874 | 1.02 (0.83,1.25) | 0.857 | 0.40 (0.04,3.56) | 0.409 | 0.898 |
| 12 | 30 | 104063861 | 104572214 | rs17034916 | 104322214 | C | T | 1.01 (0.93,1.09) | 0.836 | 1.02 (0.83,1.25) | 0.852 | 0.40 (0.04,3.56) | 0.409 | 0.889 |
| 13 | 31 | 26140990 | 26728255 | rs117674980 | 26390990 | A | G | 1.00 (0.93,1.08) | 0.906 | 1.03 (0.78,1.36) | 0.835 | - | - | 0.993 |
| 13 | 31 | 26140990 | 26728255 | rs77999054 | 26394850 | G | A | 1.01 (0.93,1.08) | 0.87 | 1.03 (0.78,1.36) | 0.848 | - | - | 0.988 |
| 13 | 31 | 26140990 | 26728255 | rs3783124 | 26458231 | C | T | 1.00 (0.93,1.08) | 0.9 | 1.06 (0.80,1.41) | 0.681 | - | - | 0.831 |
| 13 | 31 | 26140990 | 26728255 | rs74335017 | 26478255 | C | T | 1.01 (0.93,1.08) | 0.874 | 1.09 (0.82,1.46) | 0.546 | - | - | 0.723 |
| 13 | 32 | 67331127 | 67831127 | rs9571707 | 67581127 | A | G | 0.85 (0.77,0.94) | 0.002 | 1.14 (1.02,1.27) | 0.022 | 1.42 (1.11,1.81) | 0.006 | <0.001 |
| 13 | 33 | 93315090 | 93875433 | rs1932193 | 93565090 | A | C | 0.82 (0.72,0.93) | 0.003 | 1.05 (0.95,1.17) | 0.349 | 1.18 (1.01,1.38) | 0.042 | <0.001 |
| 13 | 33 | 93315090 | 93875433 | rs306675 | 93606083 | A | C | 0.82 (0.69,0.98) | 0.026 | 0.94 (0.84,1.04) | 0.238 | 1.25 (1.10,1.41) | <0.001 | <0.001 |
| 13 | 33 | 93315090 | 93875433 | rs306677 | 93613362 | C | G | 0.83 (0.69,0.98) | 0.033 | 0.94 (0.84,1.04) | 0.243 | 1.25 (1.10,1.41) | <0.001 | <0.001 |
| 13 | 33 | 93315090 | 93875433 | rs306679 | 93625433 | G | T | 0.80 (0.66,0.96) | 0.016 | 0.96 (0.86,1.07) | 0.451 | 1.23 (1.09,1.39) | <0.001 | <0.001 |
| 13 | 34 | 100252104 | 100752104 | rs72653992 | 100502104 | A | G | 0.99 (0.92,1.07) | 0.884 | 1.18 (0.90,1.55) | 0.226 | - | - | 0.15 |

| | | | | | | | | | | | | | | |
|---|---|---|---|---|---|---|---|---|---|---|---|---|---|---|
| 14 | 35 | 57055847 | 57555847 | rs12896185 | 57305847 | A | G | 0.95 (0.87,1.04) | 0.283 | 1.09 (0.96,1.23) | 0.189 | 1.18 (0.87,1.61) | 0.291 | 0.047 |
| 14 | 36 | 80321393 | 80823172 | rs1181351 | 80571393 | G | T | 0.90 (0.68,1.18) | 0.437 | 1.03 (0.92,1.16) | 0.569 | 1.00 (0.90,1.10) | 0.963 | 0.919 |
| 14 | 36 | 80321393 | 80823172 | rs28444185 | 80573172 | A | T | 0.99 (0.90,1.10) | 0.887 | 1.03 (0.92,1.16) | 0.567 | 0.89 (0.68,1.18) | 0.414 | 0.955 |
| 15 | 37 | 90077226 | 90587639 | rs28698386 | 90327226 | T | C | 1.30 (1.13,1.50) | <0.001 | 0.95 (0.86,1.05) | 0.323 | 0.88 (0.75,1.01) | 0.077 | <0.001 |
| 15 | 37 | 90077226 | 90587639 | rs6496605 | 90337639 | C | T | 1.29 (1.12,1.48) | <0.001 | 0.95 (0.86,1.05) | 0.328 | 0.87 (0.75,1.01) | 0.064 | <0.001 |
| 16 | 38 | 13669429 | 14169429 | rs75422462 | 13919429 | G | A | 0.98 (0.91,1.06) | 0.61 | 1.86 (1.30,2.67) | <0.001 | - | - | <0.001 |
| 18 | 39 | 52608233 | 53108233 | rs34352315 | 52858233 | G | A | 1.04 (0.97,1.12) | 0.273 | 0.36 (0.22,0.60) | <0.001 | - | - | <0.001 |
| 21 | 40 | 17158456 | 17658456 | rs2823575 | 17408456 | G | A | 1.00 (0.92,1.08) | 0.948 | 1.08 (0.92,1.28) | 0.328 | 0.61 (0.31,1.21) | 0.156 | 0.812 |
| 21 | 41 | 40203038 | 40703038 | rs74482819 | 40453038 | C | T | 1.00 (0.93,1.07) | 0.91 | 1.42 (0.97,2.08) | 0.073 | - | - | 0.121 |
| 22 | 42 | 18315772 | 18842083 | rs71328255 | 18565772 | A | G | 0.99 (0.92,1.06) | 0.731 | 1.67 (1.13,2.45) | 0.01 | - | - | <0.001 |
| 22 | 42 | 18315772 | 18842083 | rs71328256 | 18569926 | T | C | 0.99 (0.92,1.06) | 0.747 | 1.66 (1.13,2.45) | 0.01 | - | - | <0.001 |
| 22 | 42 | 18315772 | 18842083 | rs34680388 | 18576176 | T | A | 0.99 (0.92,1.06) | 0.743 | 1.65 (1.12,2.43) | 0.011 | - | - | 0.002 |
| 22 | 42 | 18315772 | 18842083 | rs13058445 | 18580087 | A | G | 0.99 (0.92,1.06) | 0.742 | 1.66 (1.12,2.44) | 0.011 | - | - | <0.001 |
| 22 | 42 | 18315772 | 18842083 | rs34658760 | 18584324 | A | C | 0.99 (0.92,1.06) | 0.742 | 1.66 (1.13,2.44) | 0.011 | - | - | <0.001 |
| 22 | 42 | 18315772 | 18842083 | rs34667409 | 18584379 | A | G | 0.99 (0.92,1.06) | 0.743 | 1.66 (1.13,2.44) | 0.011 | - | - | <0.001 |
| 22 | 42 | 18315772 | 18842083 | rs35137695 | 18587898 | C | T | 0.99 (0.92,1.06) | 0.744 | 1.65 (1.12,2.44) | 0.011 | - | - | 0.001 |
| 22 | 42 | 18315772 | 18842083 | rs13058179 | 18592083 | G | A | 0.99 (0.92,1.06) | 0.747 | 1.58 (1.07,2.33) | 0.021 | - | - | 0.002 |
| 22 | 43 | 29569106 | 30069106 | rs144548824 | 29819106 | G | A | 1.04 (0.97,1.12) | 0.265 | 0.52 (0.36,0.74) | <0.001 | - | - | <0.001 |

[a]The risk of fish oil supplementation for the development of Alzheimer's disease was evaluated using Cox regression models adjusted by age, sex, and 10 top genetic principal components within each SNP genotype. Genotype groups with five or fewer incident cases were not considered in the subgroup analysis.

Abbreviations: CHR, chromosome; SNP, single nucleotide polymorphism; POS, SNP position with GRCh37 assembly; REF, reference allele; ALT, alternative allele; HR, hazard ratio; CI, confidence interval; P-interaction, P value of the interaction terms.

**Table S7.** Association between fish oil supplementation and the onset of vascular dementia in each genotype of SNPs in 43 loci and *APOE* ε4 from the whole participants.

| CHR | No. Loci | Start | End | SNP | POS | REF | ALT | HR of fish oil supplementation in participants carrying REF allele[a] | | | | | | P-interaction |
|---|---|---|---|---|---|---|---|---|---|---|---|---|---|---|
| | | | | | | | | 0 | | 1 | | 2 | | |
| | | | | | | | | HR (95%CI) | P-value | HR (95%CI) | P-value | HR (95%CI) | P-value | |
| 19 | - | - | - | *APOE* ε4 | - | - | - | 0.74 (0.64,0.86) | <0.001 | 0.80 (0.68,0.95) | 0.011 | 1.21 (0.87,1.69) | 0.264 | 0.009 |
| 1 | 1 | 8027198 | 8527198 | rs116501531 | 8277198 | A | G | 0.80 (0.71,0.89) | <0.001 | 1.10 (0.59,2.05) | 0.767 | - | - | 0.413 |
| 1 | 2 | 14816263 | 15316263 | rs75837905 | 15066263 | A | G | 0.79 (0.71,0.88) | <0.001 | 1.18 (0.76,1.84) | 0.463 | - | - | 0.145 |
| 1 | 3 | 29232654 | 29732654 | rs116264291 | 29482654 | A | G | 0.78 (0.70,0.87) | <0.001 | 1.47 (0.88,2.46) | 0.138 | - | - | 0.009 |
| 1 | 4 | 101852308 | 102352308 | rs61804494 | 102102308 | G | A | 0.87 (0.77,0.98) | 0.024 | 0.69 (0.55,0.85) | <0.001 | 0.41 (0.18,0.94) | 0.035 | 0.009 |
| 1 | 5 | 106483537 | 106983795 | rs12043527 | 106733537 | G | A | 0.73 (0.65,0.82) | <0.001 | 1.13 (0.90,1.42) | 0.286 | 1.02 (0.44,2.37) | 0.964 | <0.001 |
| 1 | 5 | 106483537 | 106983795 | rs11184799 | 106733795 | G | A | 0.70 (0.62,0.80) | <0.001 | 1.09 (0.89,1.34) | 0.393 | 1.25 (0.63,2.46) | 0.529 | <0.001 |
| 1 | 6 | 110597781 | 111098352 | rs12024138 | 110847781 | C | T | 0.77 (0.69,0.86) | <0.001 | 1.25 (0.87,1.78) | 0.225 | 1.33 (0.25,7.04) | 0.739 | 0.006 |
| 1 | 6 | 110597781 | 111098352 | rs12024264 | 110848352 | C | A | 0.77 (0.69,0.86) | <0.001 | 1.25 (0.87,1.78) | 0.224 | 1.33 (0.25,7.04) | 0.739 | 0.005 |
| 1 | 7 | 222375645 | 222876659 | rs17163136 | 222625645 | T | C | 1.04 (0.88,1.22) | 0.68 | 0.72 (0.62,0.84) | <0.001 | 0.57 (0.43,0.76) | <0.001 | <0.001 |
| 1 | 7 | 222375645 | 222876659 | rs17163137 | 222626659 | T | C | 1.04 (0.88,1.23) | 0.653 | 0.72 (0.61,0.84) | <0.001 | 0.57 (0.43,0.76) | <0.001 | <0.001 |
| 2 | 8 | 100571438 | 101071438 | rs113777826 | 100821438 | C | T | 0.77 (0.69,0.86) | <0.001 | 2.16 (1.27,3.68) | 0.004 | - | - | <0.001 |
| 2 | 9 | 170757664 | 171257664 | rs4435418 | 171007664 | C | T | 0.75 (0.60,0.94) | 0.012 | 0.85 (0.73,0.99) | 0.035 | 0.83 (0.68,1.01) | 0.058 | 0.58 |
| 4 | 10 | 16312945 | 16812945 | rs148811174 | 16562945 | C | T | 0.99 (0.87,1.12) | 0.846 | 0.53 (0.44,0.65) | <0.001 | 0.48 (0.26,0.87) | 0.016 | <0.001 |
| 4 | 11 | 68145282 | 68645282 | rs149325653 | 68395282 | A | G | 0.80 (0.72,0.89) | <0.001 | 0.82 (0.39,1.71) | 0.602 | - | - | 0.84 |
| 4 | 12 | 114132769 | 114635835 | rs13123494 | 114382769 | C | A | 0.63 (0.52,0.76) | <0.001 | 0.79 (0.68,0.93) | 0.003 | 1.30 (1.02,1.66) | 0.034 | <0.001 |
| 4 | 12 | 114132769 | 114635835 | rs1525000 | 114384869 | C | T | 0.63 (0.52,0.76) | <0.001 | 0.80 (0.69,0.93) | 0.004 | 1.26 (0.99,1.61) | 0.058 | <0.001 |
| 4 | 12 | 114132769 | 114635835 | rs62314976 | 114385676 | T | C | 0.63 (0.52,0.76) | <0.001 | 0.80 (0.69,0.93) | 0.005 | 1.29 (1.01,1.65) | 0.04 | <0.001 |

| | | | | | | | | | | | | | | |
|---|---|---|---|---|---|---|---|---|---|---|---|---|---|---|
| 4 | 12 | 114132769 | 114635835 | rs13106836 | 114385835 | G | A | 0.63 (0.52,0.75) | <0.001 | 0.80 (0.69,0.93) | 0.005 | 1.29 (1.01,1.65) | 0.04 | <0.001 |
| 5 | 13 | 3009680 | 3517512 | rs7722735 | 3259680 | G | T | 0.80 (0.69,0.93) | 0.003 | 0.81 (0.69,0.96) | 0.012 | 0.78 (0.55,1.11) | 0.169 | 0.953 |
| 5 | 13 | 3009680 | 3517512 | rs59781823 | 3262724 | G | A | 0.79 (0.68,0.92) | 0.002 | 0.83 (0.70,0.97) | 0.02 | 0.74 (0.52,1.05) | 0.094 | 0.905 |
| 5 | 13 | 3009680 | 3517512 | rs60808439 | 3267512 | C | T | 0.80 (0.70,0.92) | 0.001 | 0.79 (0.66,0.95) | 0.013 | 0.90 (0.53,1.53) | 0.701 | 0.94 |
| 5 | 14 | 100147117 | 100647117 | rs2089903 | 100397117 | C | A | - | - | 2.56 (1.59,4.11) | <0.001 | 0.75 (0.68,0.84) | <0.001 | <0.001 |
| 6 | 15 | 767009 | 1267009 | rs845890 | 1017009 | C | A | 0.68 (0.59,0.78) | <0.001 | 0.98 (0.82,1.17) | 0.788 | 1.35 (0.85,2.13) | 0.205 | <0.001 |
| 6 | 16 | 115069829 | 115569829 | rs9505681 | 115319829 | C | T | 0.79 (0.71,0.88) | <0.001 | 1.31 (0.64,2.70) | 0.466 | - | - | 0.137 |
| 6 | 17 | 166801161 | 167302048 | rs6941412 | 167051161 | C | T | 0.89 (0.77,1.02) | 0.104 | 0.71 (0.60,0.85) | <0.001 | 0.64 (0.42,0.98) | 0.04 | 0.02 |
| 6 | 17 | 166801161 | 167302048 | rs12174679 | 167051368 | G | A | 0.89 (0.77,1.02) | 0.089 | 0.72 (0.60,0.85) | <0.001 | 0.62 (0.40,0.96) | 0.031 | 0.02 |
| 6 | 17 | 166801161 | 167302048 | rs963302 | 167052048 | C | G | 0.88 (0.76,1.01) | 0.079 | 0.73 (0.61,0.86) | <0.001 | 0.65 (0.43,0.97) | 0.034 | 0.033 |
| 7 | 18 | 20096169 | 20596169 | rs3114430 | 20346169 | C | A | 0.97 (0.71,1.31) | 0.826 | 0.86 (0.73,1.01) | 0.065 | 0.71 (0.61,0.84) | <0.001 | 0.069 |
| 7 | 19 | 143574310 | 144074310 | rs12670543 | 143824310 | C | A | 0.75 (0.67,0.85) | <0.001 | 1.07 (0.83,1.38) | 0.61 | 1.54 (0.48,4.97) | 0.469 | 0.009 |
| 8 | 20 | 3675912 | 4175912 | rs2042528 | 3925912 | A | T | 0.81 (0.67,0.99) | 0.039 | 0.89 (0.77,1.04) | 0.141 | 0.66 (0.53,0.84) | <0.001 | 0.341 |
| 8 | 21 | 82617116 | 83117116 | rs117216345 | 82867116 | T | A | 0.73 (0.65,0.83) | <0.001 | 1.27 (0.97,1.66) | 0.079 | 1.75 (0.70,4.39) | 0.231 | <0.001 |
| 8 | 22 | 111221071 | 111721071 | rs116871946 | 111471071 | T | C | 0.82 (0.73,0.91) | <0.001 | 0.65 (0.46,0.93) | 0.018 | 2.28 (0.43,12.17) | 0.335 | 0.714 |
| 10 | 23 | 4149849 | 4649849 | rs116876958 | 4399849 | C | T | 0.76 (0.68,0.85) | <0.001 | 1.63 (1.11,2.40) | 0.012 | - | - | <0.001 |
| 10 | 24 | 129615006 | 130115006 | rs4750683 | 129865006 | G | C | 0.78 (0.52,1.17) | 0.226 | 0.83 (0.70,0.99) | 0.036 | 0.78 (0.68,0.90) | <0.001 | 0.793 |
| 11 | 25 | 85731587 | 86233703 | rs75559794 | 85981587 | G | A | 0.79 (0.71,0.89) | <0.001 | 0.89 (0.66,1.19) | 0.422 | - | - | 0.474 |
| 11 | 25 | 85731587 | 86233703 | rs78851816 | 85983703 | G | A | 0.80 (0.71,0.89) | <0.001 | 0.85 (0.64,1.13) | 0.266 | - | - | 0.636 |
| 11 | 26 | 96280700 | 96825966 | rs754413 | 96530700 | A | G | 0.93 (0.36,2.38) | 0.88 | 0.90 (0.72,1.13) | 0.358 | 0.78 (0.69,0.87) | <0.001 | 0.28 |
| 11 | 26 | 96280700 | 96825966 | rs3018644 | 96575966 | G | A | 1.56 (0.58,4.19) | 0.376 | 0.90 (0.72,1.12) | 0.352 | 0.77 (0.68,0.87) | <0.001 | 0.075 |
| 12 | 27 | 47212480 | 47712480 | rs74523587 | 47462480 | A | C | 0.78 (0.70,0.87) | <0.001 | 1.16 (0.73,1.84) | 0.541 | - | - | 0.15 |
| 12 | 28 | 55968568 | 56468568 | rs73119275 | 56218568 | C | T | 0.78 (0.70,0.87) | <0.001 | 1.37 (0.83,2.27) | 0.221 | - | - | 0.024 |

| | | | | | | | | | | | | | | |
|---|---|---|---|---|---|---|---|---|---|---|---|---|---|---|
| 12 | 29 | 69525118 | 70042102 | rs2870901 | 69775118 | C | T | 0.57 (0.41,0.78) | <0.001 | 0.70 (0.59,0.83) | <0.001 | 0.97 (0.84,1.13) | 0.719 | <0.001 |
| 12 | 29 | 69525118 | 70042102 | rs1585705 | 69792102 | A | C | 0.53 (0.38,0.73) | <0.001 | 0.72 (0.61,0.85) | <0.001 | 0.97 (0.83,1.12) | 0.665 | <0.001 |
| 12 | 30 | 104063861 | 104572214 | rs11111842 | 104313861 | A | G | 0.75 (0.66,0.84) | <0.001 | 1.20 (0.91,1.57) | 0.192 | 4.86 (1.02,23.11) | 0.047 | <0.001 |
| 12 | 30 | 104063861 | 104572214 | rs10861145 | 104314531 | G | C | 0.75 (0.66,0.84) | <0.001 | 1.20 (0.91,1.57) | 0.193 | 4.86 (1.02,23.11) | 0.047 | <0.001 |
| 12 | 30 | 104063861 | 104572214 | rs11833702 | 104314560 | A | G | 0.74 (0.66,0.83) | <0.001 | 1.21 (0.92,1.58) | 0.176 | 4.86 (1.02,23.11) | 0.047 | <0.001 |
| 12 | 30 | 104063861 | 104572214 | rs11111843 | 104314956 | T | C | 0.75 (0.66,0.84) | <0.001 | 1.20 (0.91,1.57) | 0.193 | 4.86 (1.02,23.11) | 0.047 | <0.001 |
| 12 | 30 | 104063861 | 104572214 | rs11111844 | 104316005 | T | C | 0.75 (0.66,0.84) | <0.001 | 1.20 (0.91,1.57) | 0.191 | 4.86 (1.02,23.11) | 0.047 | <0.001 |
| 12 | 30 | 104063861 | 104572214 | rs76025677 | 104317050 | A | G | 0.75 (0.66,0.84) | <0.001 | 1.21 (0.92,1.58) | 0.174 | 4.86 (1.02,23.11) | 0.047 | <0.001 |
| 12 | 30 | 104063861 | 104572214 | rs11111846 | 104318346 | T | C | 0.75 (0.66,0.84) | <0.001 | 1.20 (0.91,1.57) | 0.191 | 4.86 (1.02,23.11) | 0.047 | <0.001 |
| 12 | 30 | 104063861 | 104572214 | rs11111849 | 104319873 | A | G | 0.75 (0.66,0.84) | <0.001 | 1.20 (0.91,1.57) | 0.192 | 4.86 (1.02,23.11) | 0.047 | <0.001 |
| 12 | 30 | 104063861 | 104572214 | rs11111850 | 104321073 | A | T | 0.75 (0.66,0.84) | <0.001 | 1.20 (0.91,1.57) | 0.191 | 4.86 (1.02,23.11) | 0.047 | <0.001 |
| 12 | 30 | 104063861 | 104572214 | rs3794246 | 104321902 | T | C | 0.75 (0.67,0.84) | <0.001 | 1.20 (0.91,1.57) | 0.194 | 4.87 (1.03,23.18) | 0.046 | <0.001 |
| 12 | 30 | 104063861 | 104572214 | rs17034916 | 104322214 | C | T | 0.75 (0.66,0.84) | <0.001 | 1.20 (0.91,1.57) | 0.191 | 4.86 (1.02,23.11) | 0.047 | <0.001 |
| 13 | 31 | 26140990 | 26728255 | rs117674980 | 26390990 | A | G | 0.85 (0.76,0.95) | 0.003 | 0.38 (0.24,0.60) | <0.001 | - | - | <0.001 |
| 13 | 31 | 26140990 | 26728255 | rs77999054 | 26394850 | G | A | 0.85 (0.76,0.95) | 0.003 | 0.38 (0.24,0.60) | <0.001 | - | - | <0.001 |
| 13 | 31 | 26140990 | 26728255 | rs3783124 | 26458231 | C | T | 0.85 (0.76,0.95) | 0.003 | 0.37 (0.23,0.59) | <0.001 | - | - | <0.001 |
| 13 | 31 | 26140990 | 26728255 | rs74335017 | 26478255 | C | T | 0.85 (0.76,0.94) | 0.003 | 0.37 (0.23,0.59) | <0.001 | - | - | <0.001 |
| 13 | 32 | 67331127 | 67831127 | rs9571707 | 67581127 | A | G | 0.78 (0.67,0.90) | <0.001 | 0.83 (0.70,0.98) | 0.028 | 0.82 (0.57,1.18) | 0.289 | 0.603 |
| 13 | 33 | 93315090 | 93875433 | rs1932193 | 93565090 | A | C | 0.78 (0.65,0.94) | 0.008 | 0.81 (0.70,0.94) | 0.007 | 0.83 (0.65,1.07) | 0.15 | 0.714 |
| 13 | 33 | 93315090 | 93875433 | rs306675 | 93606083 | A | C | 0.81 (0.63,1.03) | 0.09 | 0.81 (0.70,0.95) | 0.007 | 0.81 (0.67,0.98) | 0.027 | 0.951 |
| 13 | 33 | 93315090 | 93875433 | rs306677 | 93613362 | C | G | 0.81 (0.63,1.04) | 0.092 | 0.82 (0.70,0.95) | 0.008 | 0.81 (0.67,0.98) | 0.028 | 0.94 |
| 13 | 33 | 93315090 | 93875433 | rs306679 | 93625433 | G | T | 0.87 (0.67,1.12) | 0.281 | 0.82 (0.70,0.95) | 0.01 | 0.77 (0.64,0.93) | 0.006 | 0.42 |
| 13 | 34 | 100252104 | 100752104 | rs72653992 | 100502104 | A | G | 0.75 (0.67,0.84) | <0.001 | 1.54 (1.09,2.18) | 0.014 | 1.40 (0.27,7.27) | 0.692 | <0.001 |

| | | | | | | | | | | | | | | |
|---|---|---|---|---|---|---|---|---|---|---|---|---|---|---|
| 14 | 35 | 57055847 | 57555847 | rs12896185 | 57305847 | A | G | 0.81 (0.71,0.93) | 0.002 | 0.77 (0.64,0.93) | 0.006 | 0.87 (0.54,1.43) | 0.589 | 0.961 |
| 14 | 36 | 80321393 | 80823172 | rs1181351 | 80571393 | G | T | 1.09 (0.77,1.55) | 0.614 | 0.97 (0.82,1.14) | 0.687 | 0.65 (0.56,0.76) | <0.001 | <0.001 |
| 14 | 36 | 80321393 | 80823172 | rs28444185 | 80573172 | A | T | 0.65 (0.56,0.76) | <0.001 | 0.97 (0.82,1.13) | 0.665 | 1.10 (0.78,1.55) | 0.571 | <0.001 |
| 15 | 37 | 90077226 | 90587639 | rs28698386 | 90327226 | T | C | 0.88 (0.71,1.08) | 0.22 | 0.78 (0.67,0.91) | 0.001 | 0.81 (0.66,1.01) | 0.059 | 0.621 |
| 15 | 37 | 90077226 | 90587639 | rs6496605 | 90337639 | C | T | 0.87 (0.71,1.07) | 0.194 | 0.78 (0.67,0.90) | <0.001 | 0.78 (0.63,0.96) | 0.019 | 0.485 |
| 16 | 38 | 13669429 | 14169429 | rs75422462 | 13919429 | G | A | 0.79 (0.71,0.88) | <0.001 | 1.09 (0.64,1.84) | 0.752 | - | - | 0.292 |
| 18 | 39 | 52608233 | 53108233 | rs34352315 | 52858233 | G | A | 0.82 (0.73,0.91) | <0.001 | 0.42 (0.21,0.84) | 0.014 | - | - | 0.043 |
| 21 | 40 | 17158456 | 17658456 | rs2823575 | 17408456 | G | A | 0.70 (0.62,0.80) | <0.001 | 1.24 (1.00,1.54) | 0.053 | 0.92 (0.39,2.18) | 0.85 | <0.001 |
| 21 | 41 | 40203038 | 40703038 | rs74482819 | 40453038 | C | T | 0.76 (0.68,0.85) | <0.001 | 2.15 (1.34,3.45) | 0.002 | - | - | <0.001 |
| 22 | 42 | 18315772 | 18842083 | rs71328255 | 18565772 | A | G | 0.80 (0.72,0.89) | <0.001 | 1.04 (0.60,1.81) | 0.876 | - | - | 0.501 |
| 22 | 42 | 18315772 | 18842083 | rs71328256 | 18569926 | T | C | 0.80 (0.72,0.89) | <0.001 | 1.04 (0.60,1.81) | 0.879 | - | - | 0.503 |
| 22 | 42 | 18315772 | 18842083 | rs34680388 | 18576176 | T | A | 0.80 (0.72,0.89) | <0.001 | 1.07 (0.61,1.86) | 0.818 | - | - | 0.441 |
| 22 | 42 | 18315772 | 18842083 | rs13058445 | 18580087 | A | G | 0.80 (0.72,0.89) | <0.001 | 1.04 (0.60,1.80) | 0.884 | - | - | 0.508 |
| 22 | 42 | 18315772 | 18842083 | rs34658760 | 18584324 | A | C | 0.80 (0.72,0.89) | <0.001 | 1.07 (0.62,1.86) | 0.812 | - | - | 0.441 |
| 22 | 42 | 18315772 | 18842083 | rs34667409 | 18584379 | A | G | 0.80 (0.72,0.89) | <0.001 | 1.04 (0.60,1.81) | 0.882 | - | - | 0.506 |
| 22 | 42 | 18315772 | 18842083 | rs35137695 | 18587898 | C | T | 0.80 (0.72,0.89) | <0.001 | 1.07 (0.61,1.85) | 0.819 | - | - | 0.446 |
| 22 | 42 | 18315772 | 18842083 | rs13058179 | 18592083 | G | A | 0.79 (0.71,0.88) | <0.001 | 1.05 (0.61,1.82) | 0.861 | - | - | 0.483 |
| 22 | 43 | 29569106 | 30069106 | rs144548824 | 29819106 | G | A | 0.81 (0.73,0.90) | <0.001 | 0.73 (0.47,1.15) | 0.18 | - | - | 0.606 |

[a]The risk of fish oil supplementation for the development of vascular dementia was evaluated using Cox regression models adjusted by age, sex, and 10 top genetic principal components within each SNP genotype. Genotype groups with five or fewer incident cases were not considered in the subgroup analysis.

Abbreviations: CHR, chromosome; SNP, single nucleotide polymorphism; POS, SNP position with GRCh37 assembly; REF, reference allele; ALT, alternative allele; HR, hazard ratio; CI, confidence interval; P-interaction, P value of the interaction terms.

**Table S8.** 43 Loci interacted with fish oil supplementation in the development of dementias compared to previous discoveries.

| CHR | No. Loci | Start | End | VEP gene[a] | Closest gene[b] | Distance (bp) | Known loci[c] |
|---|---|---|---|---|---|---|---|
| 1 | 1 | 8027198 | 8527198 | - | *SLC45A1* | -100,688 | - |
| 1 | 2 | 14816263 | 15316263 | *KAZN* | *KAZN* | 0 | - |
| 1 | 3 | 29232654 | 29732654 | *SRSF4* | *SRSF4* | 0 | Omega-3% (Karjalainen et al, 2024) |
| 1 | 4 | 101852308 | 102352308 | - | *OLFM3* | -360,278 | - |
| 1 | 5 | 106483537 | 106983795 | - | *PRMT6* | -865,472 | - |
| 1 | 6 | 110597781 | 111098352 | - | *RBM15* | -32,776 | - |
| 1 | 7 | 222375645 | 222876659 | - | *HHIPL2* | -94,786 | - |
| 2 | 8 | 100571438 | 101071438 | - | *AFF3* | +62,237 | - |
| 2 | 9 | 170757664 | 171257664 | - | *MYO3B* | -26,991 | - |
| 4 | 10 | 16312945 | 16812945 | *LDB2* | *LDB2* | 0 | - |
| 4 | 11 | 68145282 | 68645282 | *CENPC* | *CENPC* | 0 | - |
| 4 | 12 | 114132769 | 114635835 | *CAMK2D* | *CAMK2D* | 0 | - |
| 5 | 13 | 3009680 | 3517512 | - | *IRX1* | -333,444 | - |
| 5 | 14 | 100147117 | 100647117 | - | *ST8SIA4* | +158,147 | - |
| 6 | 15 | 767009 | 1267009 | - | *AL033381.1* | -63,155 | - |
| 6 | 16 | 115069829 | 115569829 | - | *HS3ST5* | +655,620 | - |
| 6 | 17 | 166801161 | 167302048 | *RPS6KA2* | *RPS6KA2* | 0 | - |
| 7 | 18 | 20096169 | 20596169 | - | *ITGB8* | -24,156 | - |
| 7 | 19 | 143574310 | 144074310 | *OR2A14* | *OR2A14* | -1,841 | - |
| 8 | 20 | 3675912 | 4175912 | *CSMD1* | *CSMD1* | 0 | - |
| 8 | 21 | 82617116 | 83117116 | - | *SNX16* | +112,015 | - |
| 8 | 22 | 111221071 | 111721071 | - | *KCNV1* | +482,995 | - |
| 10 | 23 | 4149849 | 4649849 | - | *AKR1E2* | -428,972 | - |
| 10 | 24 | 129615006 | 130115006 | *PTPRE* | *PTPRE* | 0 | - |
| 11 | 25 | 85731587 | 86233703 | *EED* | *EED* | 0 | ADD (Bellenguez et al., 2022) |
| 11 | 26 | 96280700 | 96825966 | - | *JRKL* | +452,813 | - |
| 12 | 27 | 47212480 | 47712480 | - | *PCED1B* | -10,906 | - |
| 12 | 28 | 55968568 | 56468568 | *DNAJC14, ORMDL2* | *RP11-762I7.5; DNAJC14* | 0 | - |
| 12 | 29 | 69525118 | 70042102 | *YEATS4* | *YEATS4* | +38,619 | - |
| 12 | 30 | 104063861 | 104572214 | *HSP90B1* | *HSP90B1* | -9,325 | - |
| 13 | 31 | 26140990 | 26728255 | *ATP8A2, AL138815.1* | *ATP8A2* | 0 | - |
| 13 | 32 | 67331127 | 67831127 | *PCDH9* | *PCDH9* | 0 | - |
| 13 | 33 | 93315090 | 93875433 | - | *GPC6* | -273,012 | - |
| 13 | 34 | 100252104 | 100752104 | *CLYBL* | *CLYBL* | 0 | - |
| 14 | 35 | 57055847 | 57555847 | - | *OTX2* | +28,650 | - |
| 14 | 36 | 80321393 | 80823172 | - | *DIO2* | -280,928 | - |
| 15 | 37 | 90077226 | 90587639 | *ANPEP* | *MESP2* | +23,404 | - |
| 16 | 38 | 13669429 | 14169429 | - | *ERCC4* | -94,585 | - |

| | | | | | | | |
|---|---|---|---|---|---|---|---|
| 18 | 39 | 52608233 | 53108233 | - | *CCDC68* | +231,494 | - |
| 21 | 40 | 17158456 | 17658456 | - | *USP25* | +306,112 | - |
| 21 | 41 | 40203038 | 40703038 | - | *PSMG1* | -102,739 | Omega-3 PUFAs (Richardson et al, 2022; Francis et al, 2022; Borges et al, 2022; Davyson et al, 2023) Omega-6/omega-3 (Richardson et al, 2022; Borges et al, 2022) |
| 22 | 42 | 18315772 | 18842083 | *PEX26, TUBA8* | *PEX26* | 0 | - |
| 22 | 43 | 29569106 | 30069106 | *AP1B1* | *AP1B1* | 0 | - |

[a]Gene symbols were annotated by the VEP website, indicating which SNPs affected their corresponding protein-coding transcripts.

[b]Closest genes were identified as the protein-coding genes with the shortest physical distance to the top interaction signal at each locus, based on the GRCh37 genome build from the Ensembl database, and the distance was defined by the position of the top interaction signal and the start codon site of the protein-coding gene.

[c]Comparison between interaction loci and reported loci related to Alzheimer's disease and related dementias and polyunsaturated fatty acids (PUFA) traits.[3-8] No time-to-event GWAS locus of all-cause dementia, Alzheimer's disease, and vascular dementia overlapped with these 43 interaction loci.

Abbreviations: CHR, chromosome; ADD, Alzheimer's disease and related dementia; PUFA, polyunsaturated fatty acid.

**Table S9.** Association between interaction loci and dementia outcomes in the whole dataset and fish oil supplementation subgroups.

| CHR | No. Loci | Top SNP[a] | P-interaction | All[b] | | Fish oil users[b] | | Non-fish oil users[b] | |
|---|---|---|---|---|---|---|---|---|---|
| | | | | HR (95%CI) | P-value | HR (95%CI) | P-value | HR (95%CI) | P-value |
| *All-cause dementia* | | | | | | | | | |
| 1 | 2 | rs75837905 | 3.70E-05 | 1.02 (0.92,1.14) | 6.99E-01 | 1.34 (1.14,1.57) | 3.17E-04 | 0.84 (0.72,0.97) | 2.20E-02 |
| 1 | 3 | rs116264291 | 1.78E-04 | 1.16 (1.03,1.31) | 1.81E-02 | 1.51 (1.27,1.81) | **4.47E-06** | 0.95 (0.80,1.13) | 5.38E-01 |
| 1 | 4 | rs61804494 | 1.45E-04 | 0.94 (0.90,0.99) | 1.42E-02 | 0.84 (0.77,0.91) | **9.08E-06** | 1.01 (0.95,1.07) | 7.36E-01 |
| 1 | 5 | rs12043527 | 8.37E-05 | 1.00 (0.94,1.06) | 9.51E-01 | 1.15 (1.05,1.25) | 2.01E-03 | 0.91 (0.85,0.98) | 1.52E-02 |
| 1 | 6 | rs12024264 | 1.52E-06 | 1.05 (0.97,1.14) | 2.16E-01 | 1.32 (1.17,1.48) | **4.05E-06** | 0.89 (0.80,1.00) | 4.25E-02 |
| 2 | 9 | rs4435418 | 2.71E-04 | 1.05 (1.02,1.09) | 2.91E-03 | 1.14 (1.08,1.21) | **2.47E-06** | 1.00 (0.96,1.05) | 8.99E-01 |
| 6 | 16 | rs9505681 | 4.10E-05 | 1.16 (1.01,1.34) | 4.02E-02 | 1.64 (1.34,2.01) | **2.18E-06** | 0.90 (0.73,1.10) | 3.00E-01 |
| 6 | 17 | rs963302 | 3.98E-06 | 0.97 (0.93,1.01) | 1.05E-01 | 0.86 (0.81,0.92) | **3.36E-06** | 1.04 (0.99,1.09) | 1.22E-01 |
| 7 | 18 | rs3114430 | 2.09E-05 | 0.97 (0.94,1.01) | 9.17E-02 | 0.88 (0.83,0.93) | **8.95E-06** | 1.03 (0.98,1.08) | 1.92E-01 |
| 7 | 19 | rs12670543 | 3.82E-06 | 1.04 (0.98,1.10) | 1.87E-01 | 1.22 (1.12,1.33) | **6.15E-06** | 0.93 (0.86,1.00) | 6.54E-02 |
| 10 | 24 | rs4750683 | 2.44E-04 | 0.94 (0.90,0.98) | 1.24E-03 | 0.86 (0.80,0.91) | **8.99E-07** | 0.99 (0.95,1.04) | 8.12E-01 |
| 11 | 26 | rs3018644 | 6.01E-05 | 1.07 (1.02,1.13) | 9.25E-03 | 0.94 (0.87,1.02) | 1.27E-01 | 1.16 (1.09,1.24) | **7.62E-06** |
| 12 | 27 | rs74523587 | 1.54E-04 | 1.16 (1.04,1.30) | 1.05E-02 | 1.49 (1.26,1.76) | **2.41E-06** | 0.97 (0.83,1.13) | 6.71E-01 |
| 12 | 28 | rs73119275 | 6.15E-06 | 1.10 (0.97,1.24) | 1.46E-01 | 1.52 (1.27,1.81) | **3.42E-06** | 0.86 (0.72,1.02) | 8.18E-02 |
| 14 | 35 | rs12896185 | 5.71E-05 | 1.04 (1.00,1.08) | 5.07E-02 | 1.15 (1.08,1.23) | **9.16E-06** | 0.97 (0.93,1.03) | 3.16E-01 |
| 22 | 42 | rs35137695 | 6.68E-06 | 1.18 (1.04,1.34) | 1.31E-02 | 1.64 (1.37,1.96) | **6.90E-08** | 0.90 (0.74,1.08) | 2.60E-01 |
| 22 | 43 | rs144548824 | 1.83E-04 | 1.15 (1.04,1.28) | 7.85E-03 | 0.86 (0.71,1.05) | 1.30E-01 | 1.33 (1.18,1.51) | **4.89E-06** |
| *Alzheimer's disease* | | | | | | | | | |
| 1 | 1 | rs116501531 | 5.30E-05 | 1.28 (1.06,1.55) | 1.03E-02 | 1.92 (1.50,2.46) | **2.42E-07** | 0.87 (0.65,1.16) | 3.41E-01 |
| 1 | 2 | rs75837905 | 2.15E-05 | 1.17 (1.00,1.37) | 4.80E-02 | 1.68 (1.36,2.06) | **1.20E-06** | 0.84 (0.66,1.06) | 1.42E-01 |
| 1 | 6 | rs12024264 | 2.50E-05 | 1.09 (0.97,1.23) | 1.44E-01 | 1.43 (1.21,1.68) | 1.81E-05 | 0.87 (0.73,1.03) | 9.90E-02 |
| 4 | 11 | rs149325653 | 1.31E-04 | 1.28 (1.04,1.56) | 1.69E-02 | 0.65 (0.42,1.00) | 5.22E-02 | 1.70 (1.36,2.13) | **3.29E-06** |
| 5 | 13 | rs59781823 | 5.87E-05 | 1.08 (1.02,1.13) | 7.62E-03 | 0.94 (0.86,1.03) | 1.75E-01 | 1.17 (1.10,1.26) | **4.27E-06** |
| 8 | 20 | rs2042528 | 7.34E-05 | 1.07 (1.02,1.12) | 1.10E-02 | 0.94 (0.87,1.02) | 1.47E-01 | 1.16 (1.09,1.24) | **7.04E-06** |
| 8 | 22 | rs116871946 | 2.06E-04 | 1.18 (1.05,1.32) | 4.57E-03 | 0.88 (0.72,1.08) | 2.09E-01 | 1.38 (1.21,1.59) | **2.53E-06** |
| 11 | 25 | rs78851816 | 5.75E-06 | 1.12 (1.02,1.23) | 1.32E-02 | 1.42 (1.24,1.62) | **1.71E-07** | 0.93 (0.82,1.06) | 2.81E-01 |
| 13 | 32 | rs9571707 | 1.83E-06 | 1.04 (0.98,1.09) | 2.11E-01 | 1.21 (1.12,1.32) | **7.39E-06** | 0.93 (0.86,1.00) | 3.82E-02 |
| 13 | 33 | rs306675 | 1.95E-05 | 1.06 (1.00,1.11) | 4.01E-02 | 1.22 (1.12,1.32) | **2.96E-06** | 0.96 (0.90,1.03) | 2.35E-01 |
| 15 | 37 | rs28698386 | 1.20E-04 | 0.94 (0.89,0.99) | 1.72E-02 | 0.83 (0.77,0.90) | **5.96E-06** | 1.02 (0.96,1.09) | 5.26E-01 |
| 16 | 38 | rs75422462 | 2.28E-04 | 1.26 (1.05,1.50) | 1.17E-02 | 1.78 (1.40,2.26) | **1.98E-06** | 0.91 (0.69,1.19) | 4.94E-01 |

| | | | | | | | | | |
|---|---|---|---|---|---|---|---|---|---|
| 18 | 39 | rs34352315 | 3.42E-05 | 1.36 (1.11,1.66) | 2.87E-03 | 0.63 (0.40,1.00) | 4.86E-02 | 1.88 (1.50,2.35) | **4.44E-08** |
| 22 | 43 | rs144548824 | 2.06E-04 | 1.09 (0.93,1.28) | 2.81E-01 | 0.69 (0.50,0.94) | 2.02E-02 | 1.37 (1.14,1.65) | 8.93E-04 |
| *Vascular dementia* | | | | | | | | | |
| 1 | 5 | rs11184799 | 5.86E-05 | 1.15 (1.04,1.27) | 7.08E-03 | 1.49 (1.27,1.74) | **6.05E-07** | 0.98 (0.86,1.11) | 7.49E-01 |
| 1 | 7 | rs17163137 | 3.82E-05 | 0.96 (0.89,1.03) | 2.35E-01 | 0.77 (0.68,0.88) | 6.29E-05 | 1.07 (0.98,1.17) | 1.44E-01 |
| 2 | 8 | rs113777826 | 1.20E-04 | 1.37 (1.05,1.78) | 1.83E-02 | 2.39 (1.70,3.37) | **5.37E-07** | 0.84 (0.56,1.27) | 4.06E-01 |
| 4 | 10 | rs148811174 | 6.38E-08 | 1.07 (0.98,1.17) | 1.16E-01 | 0.73 (0.62,0.87) | 3.35E-04 | 1.27 (1.14,1.41) | **6.37E-06** |
| 4 | 12 | rs13106836 | 5.65E-06 | 1.06 (0.99,1.14) | 1.19E-01 | 1.33 (1.18,1.50) | **3.58E-06** | 0.94 (0.85,1.02) | 1.45E-01 |
| 5 | 14 | rs2089903 | 7.41E-07 | 0.86 (0.68,1.08) | 1.84E-01 | 0.46 (0.34,0.61) | **1.44E-07** | 1.52 (1.05,2.21) | 2.67E-02 |
| 6 | 15 | rs845890 | 1.49E-04 | 1.13 (1.04,1.23) | 5.29E-03 | 1.40 (1.22,1.61) | **1.81E-06** | 1.00 (0.89,1.11) | 9.56E-01 |
| 8 | 21 | rs117216345 | 2.14E-05 | 1.14 (1.00,1.29) | 4.75E-02 | 1.59 (1.32,1.93) | **1.67E-06** | 0.91 (0.76,1.08) | 2.82E-01 |
| 10 | 23 | rs116876958 | 1.65E-04 | 1.33 (1.10,1.62) | 3.05E-03 | 2.03 (1.55,2.65) | **2.28E-07** | 0.98 (0.74,1.29) | 8.63E-01 |
| 12 | 29 | rs1585705 | 6.05E-05 | 0.91 (0.84,0.98) | 1.26E-02 | 1.13 (0.99,1.29) | 6.60E-02 | 0.81 (0.74,0.89) | **8.51E-06** |
| 12 | 30 | rs11833702 | 1.96E-04 | 1.19 (1.04,1.36) | 1.24E-02 | 1.63 (1.33,1.99) | **3.04E-06** | 0.97 (0.81,1.16) | 7.44E-01 |
| 13 | 31 | rs3783124 | 2.08E-04 | 1.28 (1.06,1.53) | 8.79E-03 | 0.67 (0.44,1.02) | 5.91E-02 | 1.61 (1.32,1.98) | **4.15E-06** |
| 13 | 34 | rs72653992 | 5.78E-05 | 1.37 (1.16,1.62) | 2.26E-04 | 2.06 (1.62,2.61) | **2.88E-09** | 1.02 (0.80,1.29) | 8.76E-01 |
| 14 | 36 | rs28444185 | 8.16E-05 | 1.09 (1.01,1.18) | 2.59E-02 | 1.34 (1.18,1.52) | **5.21E-06** | 0.97 (0.88,1.07) | 5.56E-01 |
| 21 | 40 | rs2823575 | 4.40E-05 | 1.11 (1.00,1.24) | 5.99E-02 | 1.47 (1.24,1.73) | **6.02E-06** | 0.93 (0.80,1.07) | 3.16E-01 |
| 21 | 41 | rs74482819 | 1.74E-05 | 1.25 (0.99,1.58) | 6.56E-02 | 2.18 (1.61,2.97) | **6.10E-07** | 0.76 (0.53,1.10) | 1.46E-01 |

[a]The most significant interaction SNPs in loci for each outcome.

[b]The association between interaction signals and dementia outcomes was evaluated by Cox regression models adjusted by age, sex, and 10 top genetic principal components.

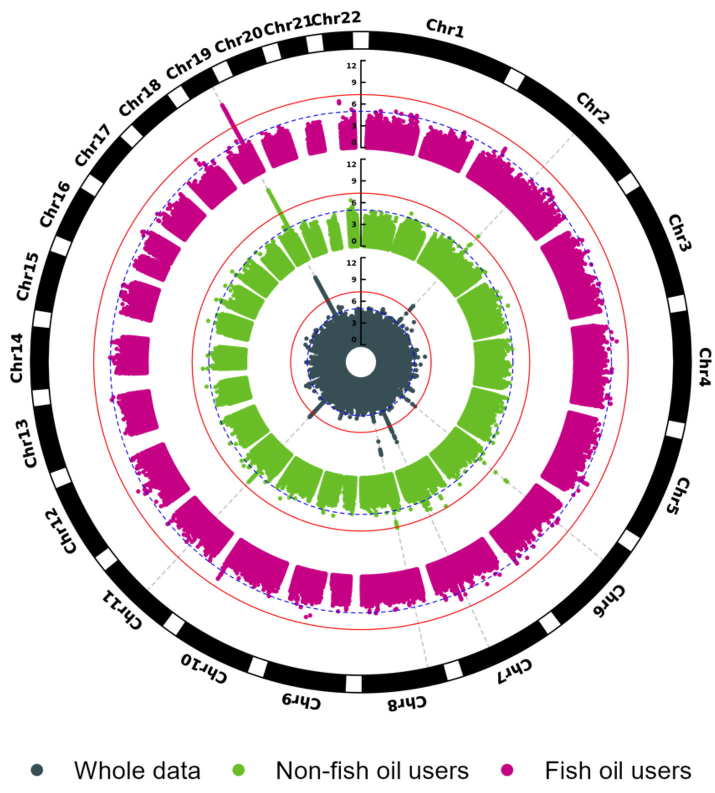

**Figure S1**. Manhattan plots of GWAS of all-cause dementia by fish oil supplementation

Circular Manhattan plot for time-to-event GWAS of all-cause dementia in whole data (dark cyan), non-fish oil users (light green), and fish oil users (purple). Variants with a -log10(p-value) below 11 are not shown. Thresholds of genome-wide significance (p-value < 5e-8) and suggestive significance (p-value < 1e-5) were shown by a solid red line and dashed blue line, respectively. The positions of genome-wide significant signals were demonstrated by cross-circle grey dashed lines.

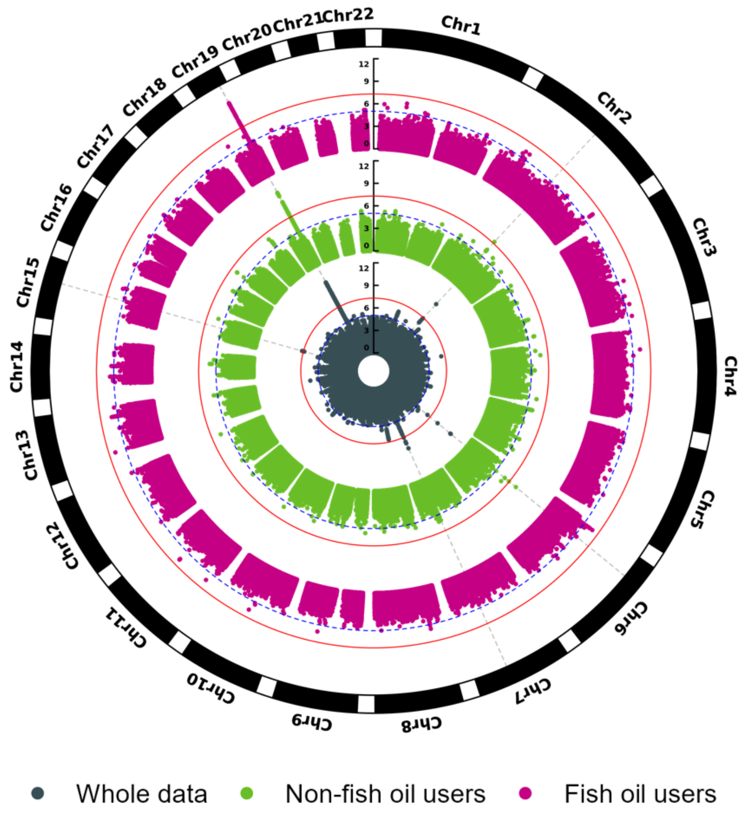

**Figure S2.** Manhattan plots of GWAS of Alzheimer's disease by fish oil supplementation

Circular Manhattan plot for time-to-event GWAS of Alzheimer's disease in whole data (dark cyan), non-fish oil users (light green), and fish oil users (purple). Variants with a -log10(p-value) below 11 are not shown. Thresholds of genome-wide significance (p-value < 5e-8) and suggestive significance (p-value < 1e-5) were shown by a solid red line and dashed blue line, respectively. The positions of genome-wide significant signals were demonstrated by cross-circle grey dashed lines.

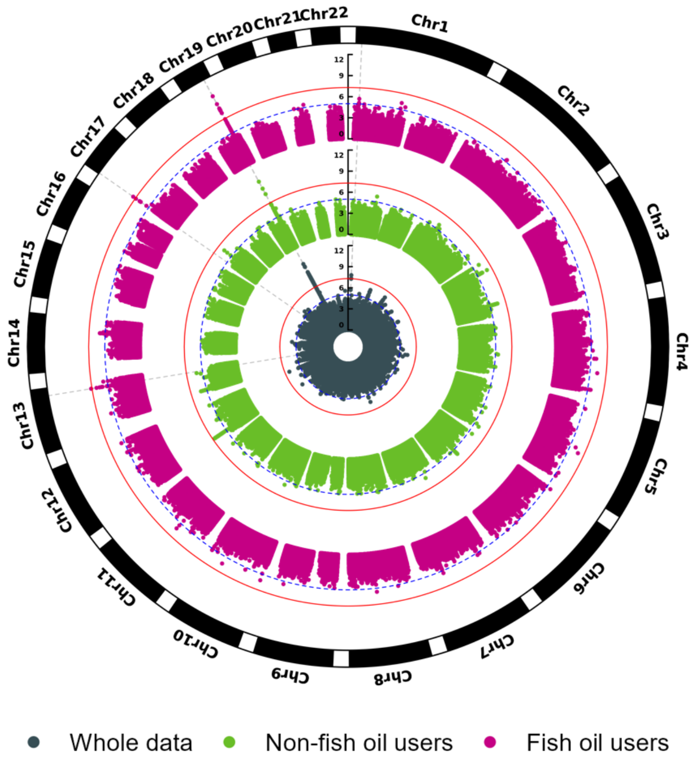

**Figure S3**. Manhattan plots of GWAS of vascular dementia by fish oil supplementation

Circular Manhattan plot for time-to-event GWAS of vascular dementia in whole data (dark cyan), non-fish oil users (light green), and fish oil users (purple). Variants with a -log10(p-value) below 11 are not shown. Thresholds of genome-wide significance (p-value < 5e-8) and suggestive significance (p-value < 1e-5) were shown by a solid red line and dashed blue line, respectively. The positions of genome-wide significant signals were demonstrated by cross-circle grey dashed lines.

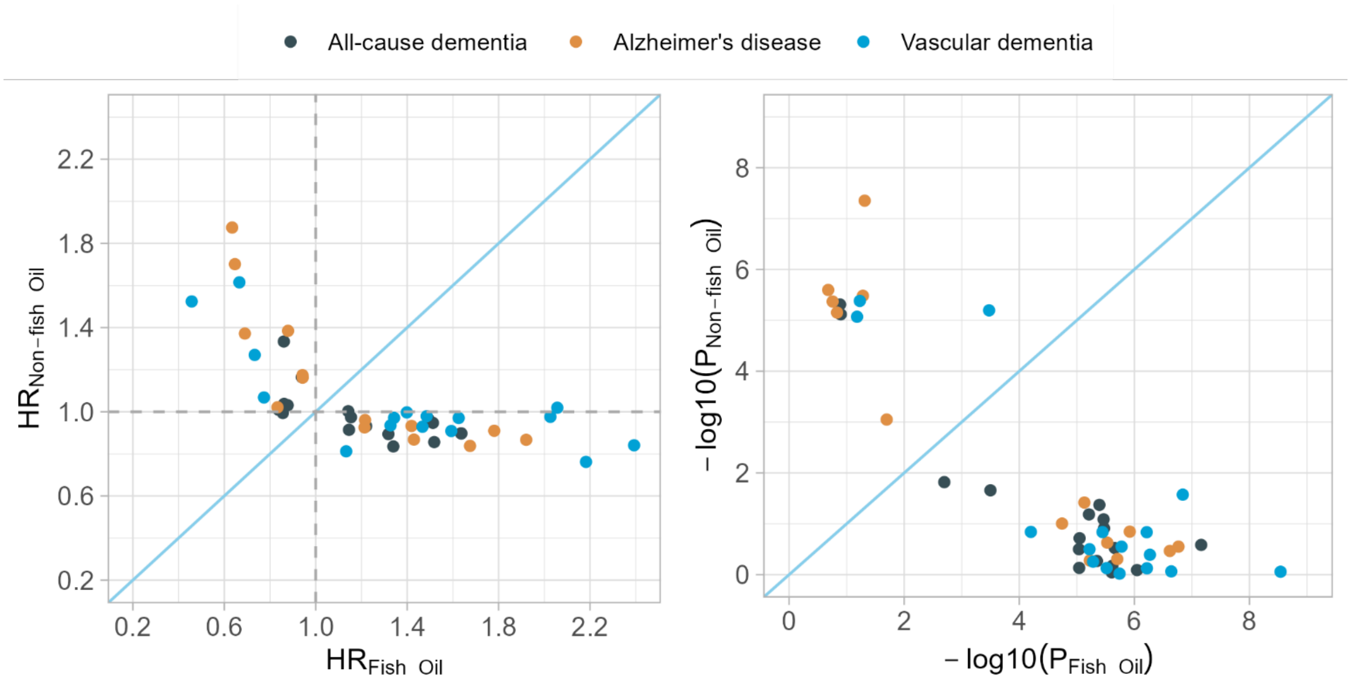

**Figure S4**. Comparison of the associations between interaction loci and dementia outcomes stratified by fish oil supplementation.

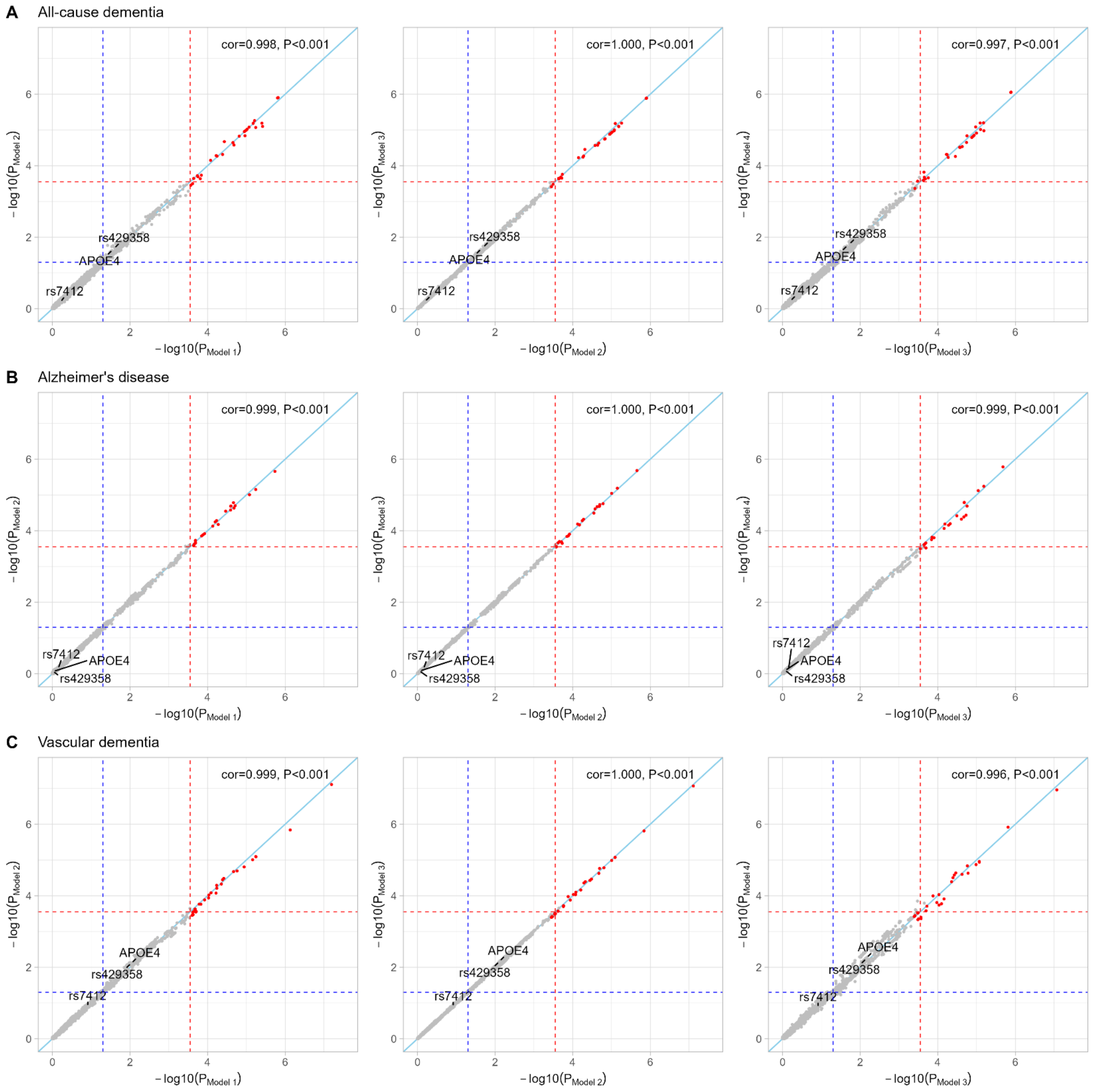

**Figure S5.** Interaction analysis of candidate SNPs with fish oil supplementation in the development of dementia using Model 1-4.

A) Comparison of P values for SNP × fish oil interaction in the development of all-cause dementia between Model 1 and Model 2, Model 2 and Model 3, and Model 3 and Model 4, respectively. B) Comparison of p-values for SNP × fish oil interaction in the development of Alzheimer's disease between Model 1 and Model 2, Model 2 and Model 3, and Model 3 and Model 4, respectively. C) Comparison of p-values for SNP × fish oil interaction in the development of vascular dementia between Model 1 and Model 2, Model 2 and Model 3, and Model 3 and Model 4, respectively. The SNPs that significantly interacted with fish oil intake status in Model 1 were demonstrated by red points. The red dashed line and blue dashed line referred to the Bonferroni correction threshold (p-value=2.8e-4) and nominated threshold (p-value=0.05), respectively. Pearson tests were conducted to assess the relatedness of the p-values between 2 datasets.

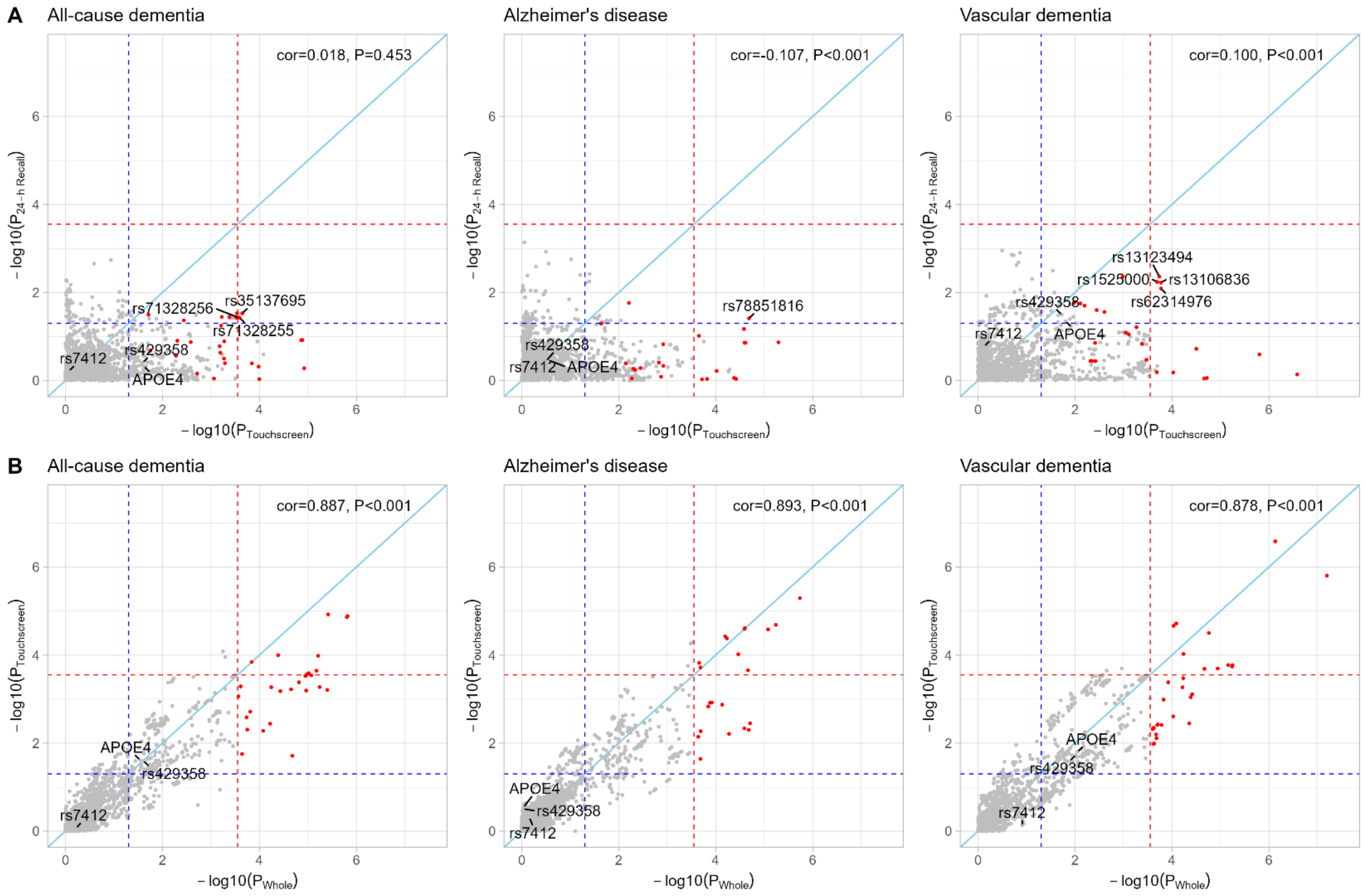

**Figure S6.** Interaction analysis of candidate SNPs with fish oil supplementation from the touchscreen questionnaire and 24-hour recall questionnaire in the development of dementia by model 1.

A) Comparison of p-values for SNP × fish oil interaction inferred from exclusive touchscreen questionnaire (N=201,627) and 24-hour recall questionnaire (N=156,004). B) Comparison of p-values for SNP × fish oil interaction between the whole dataset (N=357,631) and exclusive touchscreen questionnaire (N=201,627). SNPs that significantly interacted with fish oil intake status in the whole dataset were demonstrated by red points. The Red dashed line and blue dashed line referred to the Bonferroni correction threshold (p-value=2.8e-4) and nominated threshold (p-value=0.05), respectively. Pearson tests were conducted to assess the relatedness of the p-values between 2 datasets.

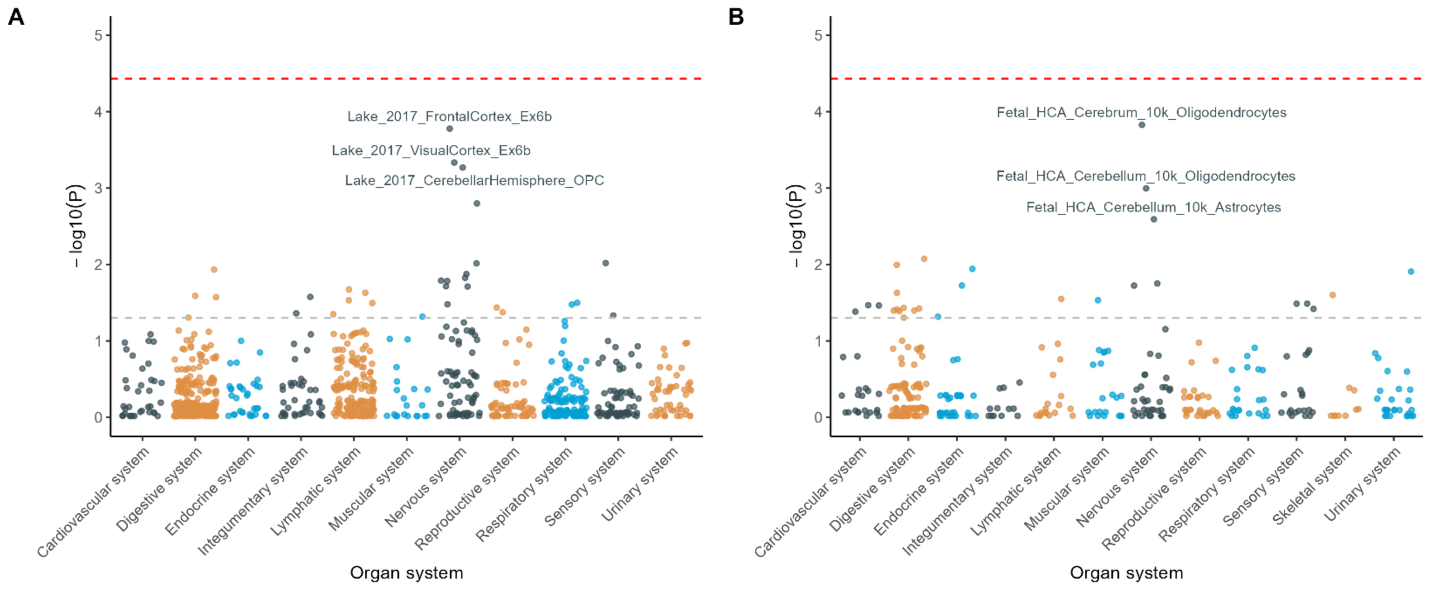

**Figure S7.** Protein-coding genes enrich in tissues by adult and fetal status.

A) Tissue-cell types of genes specifically expressed across 941 adult organ systems. B) Tissue-cell types of genes specifically expressed across 364 fetal organ systems. P-values of cell-type-specific enrichment analysis after normalizing the effect of gene set length by permutation-based method. Red and grey dashed lines represent thresholds for Bonferroni-corrected significance (0.05/total number of tissue-cell types) and nominal significance (0.05), separately.
